# Early life experiences are associated with later life DNA methylation signatures in the Health and Retirement Study

**DOI:** 10.64898/2026.07.24.26358676

**Authors:** Nita Kanney, Scarlet Cockell, Herong Wang, Mingzhou Fu, John Dou, Margaret T. Hicken, Devon Payne-Sturges, Belinda L. Needham, Erin B. Ware, Kelly M. Bakulski

## Abstract

Long term associations of early life experiences with later life DNA methylation are understudied. In the U.S. Health and Retirement Study, participants self-reported early life experiences, including years in school, smoking during childhood, growing up in a rural area, and living with a grandparent. Later life DNA methylation was measured in blood for participants with a mean age of 69.6 years at 731,474 sites. We tested for associations between each early life experience with DNA methylation age acceleration, global and site-specific methylation, and enriched biological pathways. We compared results across early life experiences. Participants (N = 3,562) were 58.8% female and 68.1% non-Hispanic White. They reported 13 mean years in school, 18.3% smoked during childhood, 42.7% grew up in a rural area, and 26.8% lived with a grandparent. Fewer years in school (0.10, 95% CI: 0.06, 0.14) and smoking during childhood (0.65, 95% CI: 0.32, 0.97) were associated with accelerated GrimAge in later life, while living in a rural area and living with a grandparent were not associated. Early life experiences were associated (p<1×10^-4^) with distinct DNA methylation sites, specifically 574 sites for years in school, 20 for smoking during childhood, 49 for growing up in a rural area, and 23 for living with a grandparent. For example, one fewer year in school was associated with 0.47 (p-value = 4.61×10^-15^) lower percent methylation at cg07318158 in *OTUD7B*. Sites associated with our early life exposures were enriched for unique pathways. Years in school was enriched for embryonic development and cell structure pathways, smoking during childhood was enriched for nervous system development, exocytosis, and cell adhesion and structure pathways, growing up in a rural area was enriched for cell and vesicle processing pathways, and living with a grandparent was enriched for hormone regulation and protein breakdown pathways. Findings suggest our early life exposures are associated with unique DNA methylation patterns in later life, which can potentially allow for separate biomarker opportunities aimed at early intervention of adverse later life outcomes associated with these exposures.

## Introduction

The early life period spans from birth to adolescence and is considered a sensitive window of susceptibility. Experiences during this period, such as growing up in a rural area, educational attainment, living with a grandparent, and cigarette smoking behaviors, shape adulthood socioeconomic and aging trajectories.^1,2^ These exposures can occur independently, or individuals can experience more than one of these exposures during early life, and exposure to one factor can increase the likelihood of experiencing a second factor. For example, growing up in a rural area was associated with lower educational attainment, which jointly increased the risk for Alzheimer’s disease in later life.^3^ Childhood rural residence was also associated with lower financial assets in young adulthood,^4^ an earlier age of adolescent smoking initiation,^5^ and increased risk of daily smoking^6^ compared to urban adolescents. Additionally, living with a grandparent in early life was associated with potential financial hardships,^7^ higher parental educational attainment,^8^ and lower personal educational attainment.^9,10^ Findings suggest early life exposures can become biologically embedded, influencing physiological processes across the life course. One mechanism that may connect early life and adult health is DNA methylation.^11,12^

DNA methylation is an epigenetic modification where a methyl group is added to a cytosine base along a DNA strand and can subsequently influence gene expression.^13^ This modification is not permanent and is sensitive to various social and environmental exposures. However, the timing, duration, and impact of these exposures on DNA methylation is not fully characterized. Prior studies have examined individual dimensions of DNA methylation with social and environmental exposures, including DNA methylation age (chronologic clocks: Horvath,^14^ Hannum^15^ and phenotypic clocks: GrimAge,^16^ MPOA,^17^ Levine^18^), global methylation, and site-specific DNA methylation. Socio-environmentally sensitive exposures may affect later life DNA methylation, but it is not fully understood how these past exposures, individually and in combination, impact multiple dimensions of DNA methylation in the same older adult population.

Self-reported early life exposures, such as socioeconomic status and smoking, have been associated with measures of DNA methylation in midlife and older adulthood.^19–21^ Among the different DNA methylation age measures tested, less education is consistently suggestively linked to higher GrimAge and MPOA DNAm age acceleration among middle and older adults.^19,20^ Smoking in early life was associated with accelerated GrimAge independent of later life smoking behaviors.^21^ However, findings across studies are conflicting in the magnitude of the associations across studies, and there is inconsistency in the age acceleration measures included. Studies examining early life exposures and site-specific methylation have focused on maternal educational attainment, adolescent smoking, and adult smoking.^22–26^ Maternal educational attainment^22^ and cigarette use in adulthood^25^ were significantly associated (FDR adjusted p-value<0.05) with DNA methylation signatures. Few studies have explored whether an individual reported growing up in a rural area and/or living with a grandparent, with multiple dimensions of DNA methylation. These experiences can co-occur; thus, it is important to examine exposures individually and in combination. Furthermore, exploration of multiple types of early life exposures on later life differences in DNA methylation is important for understanding the shared and singular pathways that may be intervenable, on which we may base public health interventions.

To better understand whether different early life exposures have overlapping or unique DNA methylation signatures that persist until older adulthood, our study will use the long-running U.S. Health and Retirement Study. We will focus on four early life exposures: growing up in a rural area, years in school, living with a grandparent, and smoking during childhood. We hypothesize these four early life exposures will be significantly associated with later life differences in DNA methylation, measured using age acceleration, global methylation, site-specific methylation, and enriched biological pathways. To understand whether these DNA methylation sites are unique across our exposures, we will examine the consistency of these findings through replication with previously identified sites. Findings from this study will characterize the specific and shared impacts of modifiable early life exposures on DNA methylation in older adults.

## Materials and Methods

### Study population

The Health and Retirement Study (HRS) is a national longitudinal study that began in 1992 (sponsored by the National Institute on Aging grant U01 AG009740).^27^ Using a multi-stage probability sampling design, the HRS biannually interviews individuals in the U.S. who are 51 years and older on various topics across their life course. The HRS replenishes the sample every 6 years, including oversampling of underrepresented racial/ethnic groups. Survey questions capture experiences related to health, economic status, and social conditions. The surveys also retrospectively ascertained respondents’ early life experiences. Participants provided written informed consent at the time of participation. These secondary data analyses are approved by the Institutional Review Board of the University of Michigan (HUM00128220). For this study, we use a retrospective longitudinal study design, where early life conditions were queried between 2008 and 2016, and DNA methylation was measured in 2016. Our study accessed publicly available data^28^, sensitive health data^29^, and restricted DNA methylation data^30^ (Accession Number: NG00153.v1).

### Early life exposure measures

We defined the early life period as childhood and adolescence prior to age 17. Our baseline for capturing early life experiences starts in 2008 because smoking during childhood was not asked on core surveys until 2008. Thus, we included early life responses retrospectively collected in core surveys between 2008 and 2016. If a participant did not complete this survey at their time of enrollment, they were asked at a subsequent interview. The HRS aggregated responses to these questions across core surveys, including performing data cleaning steps to remove secondary responses in the rare case a participant was asked twice.^31^

Specific early life exposure survey questions are provided in **Supplemental Table 1**. All early life exposure variables are modeled such that a more deleterious event is the exposure (e.g., smoking during childhood vs. not smoking during childhood). We evaluated four early life exposures: growing up in a rural area, years in school, living with a grandparent, and childhood smoking. Participants were asked whether they were living in a rural area most of the time when they were age 10 (yes, no=reference). Participants self-reported the number of years of school they completed (0 to 17 years). We examined whether a participant reported living in the same household with a grandparent for a year or more before they were 17 (yes, no=reference). Finally, we assessed early life smoking as whether a participant regularly smoked cigarettes when they were in grade or high school (yes, no=reference).

### DNA methylation assay and data processing

In 2016, the HRS asked a nonrandom subset of participants to provide whole blood samples for DNA extraction and assay on the Infinium Methylation EPIC (v1.0) BeadChip array at the University of Minnesota’s Advanced Research and Diagnostic Lab.^32,33^ Sample processing and HRS quality control (QC) measures have been provided elsewhere.^33^ QC sample checks included adequate detection, potential sex-mismatch, and blinded duplicates with low correlation conducted in the *minfi* package.^34^ Sample plate was noted as a technical covariate. This resulted in a sample of 4,018 participants with DNA methylation data released by the HRS.

Our team downloaded the DNA methylation image files for these samples from the National Institute on Aging Genetics of Alzheimer’s Disease Data Storage Site (NIAGADS) (https://dss.niagads.org/datasets/ng00153/; Accession Number: NG00153.v1) and carried out a second set of QC steps^35^ in the *ewastools* package.^36^ Probes were excluded for poor detection (p>0.0001, 95,491 sites) and cross-reactivity (38,904 sites).^37^ Samples were excluded for low intensity and failed control probe checks (n=10), unexpected high genetic matching (n=18), mismatched genotype data (n=5), and granulocyte proportions that were not within standard ranges (n=5). We performed noob background and dye-bias correction,^38^ followed by BMIQ normalization.^39^ There were 3,980 participants with eligible DNA methylation data at 731,696 sites. Using the Salas reference panel^40^ and Houseman method,^41^ we estimated cell type proportions (granulocytes, natural killer cells, B cells, CD4+ T cells, CD8+ T cells, and monocytes).

### DNA methylation age acceleration and global DNA methylation

DNA methylation clocks are summary measures of specific sites strongly correlated with chronological age and phenotypic age, allowing examination of differences in an individual’s biological age relative to their calendar age.^42^ DNA methylation clocks were provided by the HRS.^43^ Phenotypic clocks incorporate behavioral factors and aging-related health outcomes in their prediction of biological aging. Due to their methodological rigor and strong association with various health outcomes, our primary measures were three phenotypic clocks: GrimAge,^16^ MPOA (DunedinPoAm38),^17^ and Levine (PhenoAge).^18^ For comparability across clocks, MPOA was converted from a rate to years by multiplying by a participant’s current calendar age by their MPOA age. Although chronologic clocks only reflect chronological age, we evaluated the Horvath^14^ and Hannum^15^ clocks as sensitivity measures. To capture biological aging among our sample, we separately regressed calendar age onto each of the clocks and extracted the residuals, which we used as our measure of age acceleration. A DNA methylation age acceleration value greater than zero indicated accelerated biologic aging, while a value of zero or below was not accelerated. An example scatterplot and regressed effect of calendar age on GrimAge DNA methylation age to calculate age acceleration is displayed in **Supplemental Figure 1**. Discordance between an individual’s calendar and biological age, particularly when it results in accelerated aging, is associated with adverse outcomes, like cognitive function, chronic disease, and mortality.^44,45^

Global methylation captures the average level of DNA methylation within our samples across different regions of the genome (island, open sea, shelf, and shore). This allows for a broader understanding of DNA methylation patterns. To annotate each site to a region, we used the IlluminaHumanMethylationEPICanno.ilm10b4.hg19 package^46^ and calculated the average DNA methylation level across all sites in a given region.

### Covariate measures

Participants’ self-reported demographic factors, including sex (female, male=reference), and race/ethnicity (non-Hispanic Black, Hispanic, non-Hispanic White=reference). Participants provided their birthdate at enrollment, and we calculated their calendar age in 2016 (time of DNA methylation measure). Adult health, socioeconomic, and behavior variables were queried by the HRS every two years. We selected a participant’s most recent response between 2008 and 2016 for model adjustment. Adult health factors were obtained from the RAND HRS Longitudinal File 2018 (V1), provided by the RAND Center for the Study of Aging. Participants self-reported whether they ever had high blood pressure (yes, no=reference), ever had diabetes (yes, no=reference), and the amount of vigorous physical activity they participated in (never, 1 per week through 1-3 per month, every day through >1 per week=reference). Although these variables may act as potential mediators and/or confounders, in this study, we will assess these health covariates as precision variables based on their prior associations with differences in DNA methylation.^47–50^ We also considered the participants’ socioeconomic and behavioral factors. We assessed participants’ current adult urban-rural residence status. County of residence at the time of interview was linked to the United States Department of Agriculture’s Rural-Urban Continuum Codes (suburban, ex-urban, urban=reference), which are defined using 2010 Census data.^51^ We adjusted our models for self-reported parental education (less than high school, high school & beyond=reference). We also accounted for the participant’s current, adult median total of all household assets. This variable was calculated as a summation of a participant’s wealth components minus all debts they hold. Smoking behaviors can impact DNA methylation; thus, we accounted for the participant’s current, adult smoking status (current, former, never=reference). Participants who reported smoking during early life but never smoking as an adult (n=7) were excluded. Adjusting for self-reported health, socioeconomic, and behavior variables may attenuate the above associations due to the impact of later life factors on the associations between early life exposures and later life DNA methylation.

### Statistical analysis

Analyses were conducted in R statistical software (version 4.5.2).^52^ Code to conduct analyses is available on GitHub (https://github.com/bakulskilab/EarlyLife_DNAm_HRS). Participants were included in the analytic sample if they had complete information on DNA methylation, early life exposures, and covariate measures. We visualized participant exclusions using a flowchart (**Figure 1**). Further, we calculated the distributions of continuous variables using mean and standard deviation, and categorical variables using count and frequency within the included and excluded samples. We assessed the included sample for potential bias by comparing it to the excluded sample using the Wilcoxon rank sum test for continuous variables and Fisher’s exact test for categorical variables.

**Figure 1.**
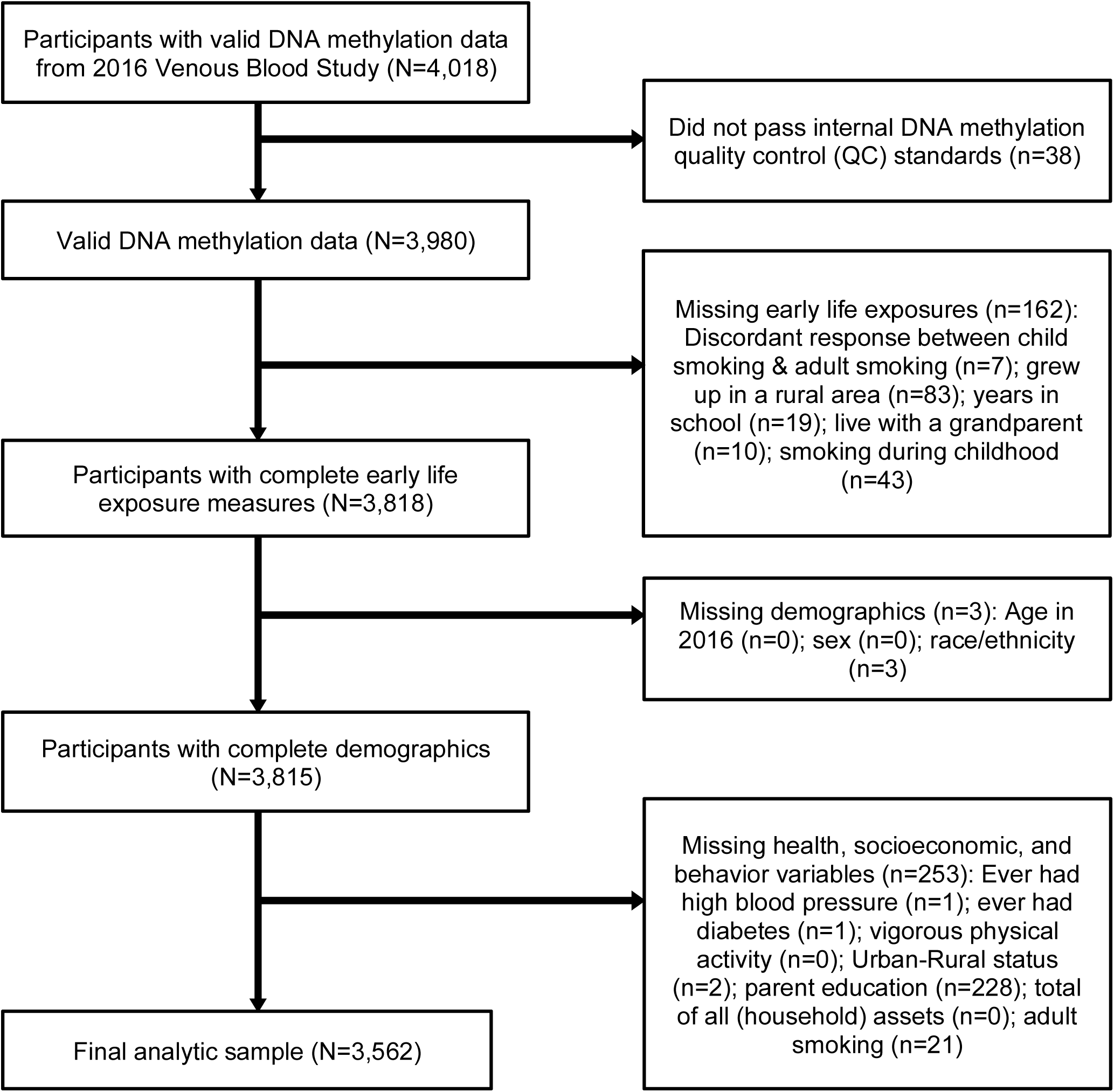
Inclusion and exclusion flow chart of participants from the U.S. Health and Retirement Study with available DNA methylation data from the 2016 Venous Blood Study (N=3,562) Inclusion and exclusion flow chart of participants from the U.S. Health and Retirement Study with available DNAm data from the 2016 Venous Blood Study. Boxes on the right describe specific variables from our analysis with missing data. N represents the number of participants included, while n represents the number of participants excluded from our analysis.

Within our included sample, we examined bivariate distributions of covariates by each of the four early life exposures and GrimAge acceleration. We stratified by a dichotomized GrimAge acceleration residual, a well validated estimate of biological aging, due to prior findings of its sensitivity to various early life, health, and socioeconomic factors, particularly in older populations.^53,54^ We visualized distributions of GrimAge acceleration by early life exposure status using raincloud plots. We assessed correlation among all DNA methylation age acceleration measures, calendar age, methylation by molecular location, cell type composition, years in school, and household assets through a Pearson correlation matrix. We examined correlations between categorical variables using Cramer’s V correlation matrix.

To understand whether our four early life exposures were associated with accelerated DNA methylation aging, we ran separate unadjusted, demographic, health, and socioeconomic and behavior adjusted linear regression models for each combination of early life exposure and DNA methylation age acceleration measure. Demographic models were adjusted for age, sex, race/ethnicity, cell type composition (Granulocytes, natural killer cells, B cells, CD4+ T cells, and CD8+ T cells), and sample plate. Monocytes were left out of the regression models due to multicollinearity issues. Health models were further adjusted for ever having high blood pressure, ever having diabetes, and self-reported vigorous physical activity. Socioeconomic and behavior models were further adjusted for adult urban-rural status, parent education, total of all household assets, and adult smoking status. We reported the beta and 95% confidence intervals (CI), representing the adjusted difference in DNA methylation age acceleration in years. We visualized the estimates using forest plots and assessed potential multicollinearity of covariates in our regression models using the Variance Inflation Factor (VIF).^55^

We used the same modeling framework for our global methylation outcome measures. We ran additional unadjusted and multivariable linear regression models for each early life exposure with each molecular location’s global methylation as described above. The effect estimates represent the adjusted difference in mean methylation at each molecular location (significance threshold of adjusted p-value<0.05).

Site-specific methylation allows for a deeper exploration of the sites, genes, and pathways sensitive to our exposures.^56^ We conducted an epigenome-wide association study (EWAS) to estimate site-specific associations with early life exposures. For our EWAS, we ran the same multivariable linear regression models as our age acceleration and global methylation measures for each early life exposure variable and each DNA methylation site using the *limma* package.^57^ We created diagnostic quantile-quantile (QQ) plots with lambda values to assess inflation in site-specific model results. We reported the effect estimates, which represent the adjusted difference in percent DNA methylation and are visualized with volcano plots. We compared effect estimates across each early life exposure for our socioeconomic and behavior model results using scatterplots and Pearson correlation. The Benjamini-Hochberg method was applied using the false discovery rate (FDR) adjusted p-values to account for multiple comparisons.^58^ FDR significance in our study was designated as an adjusted p-value<0.05. We set a nominal significance level of p<1×10^-4^ for tested sites.

We sought to understand the association between each exposure and outcome, controlling for the other three exposures. Thus, we ran a mixtures model, adjusted for the same covariates in the socioeconomic and behavior models, for age acceleration, global methylation, and site-specific methylation, with each early life exposure variable included in the same model. *Replication and additional analyses*

Though no EWAS currently exist using the same outcomes from early life we explore in our analyses, we assessed replication by comparing our socioeconomic and behavior adjusted EWAS models with two existing EWAS similar in outcome. In the Pregnancy and Childhood Epigenetics (PACE) consortium, Choudhary et al.^22^ conducted a meta-analysis across 37 cohorts examining EWAS between maternal educational attainment and their child’s DNA methylation levels across three developmental periods (i.e., 27 studies at birth, 6 studies at childhood, 4 studies at adolescence), adjusting for sex, cell type proportions, and cohort. Samples from the included studies were obtained from cord and peripheral blood. We examined the provided summary statistics from 476 sites for overlap with the top sites from our EWAS for years in school. In the CHARGE consortium, Joehanes et al.^25^ conducted a meta-analysis of 16 cohorts with EWAS of adult smoking status (former, current, never). The prior studies adjusted for age, sex, cell type proportions, and cohort and obtained DNA methylation levels from whole blood, buffy coat, and CD14+ samples. For former smoking, 2,568 sites were available and for current smoking, summary statistics were available for the top 18,760 sites. Comparisons of our EWAS results with these prior studies include scatterplots, Pearson correlations, and an UpSet plot of overlapping top hits (p<1×10^-4^). Our EWAS on growing up in a rural area and living with a grandparent are the first to our knowledge, and we therefore do not include EWAS comparison for this outcome.

To understand the biological processes underlying the findings of our EWAS, we conducted a gene set enrichment analysis using *methylGSA*.^59^ We input all sites from our socioeconomic and behavior models separately for each exposure into the methylRRA function. This function allows for p-value adjustment for multiple sites associated with a single gene from a preranked gene set enrichment analysis (GSEAPreranked approach) without prior filtering for significant sites or setting a cutoff, which limits potential false positives.

## Results

### Sample characteristics

After excluding participants due to missing information on exposures and covariates, our analytic sample had 3,562 participants (**Figure 1**). Excluded participants were older biologically, had fewer years of schooling, were more likely to be current smokers, and had lower median total household assets relative to the included sample (**Supplemental Table 2**). The analytic sample was on average 69.6 years old, predominantly female, and non-Hispanic White (**Table 1**). A majority of the analytic sample reported not growing up in a rural area (57.3%), not living with a grandparent (73.2%), not smoking during childhood (81.7%), and having a mean of 13 years in school (**Table 1**).

**Table 1.**
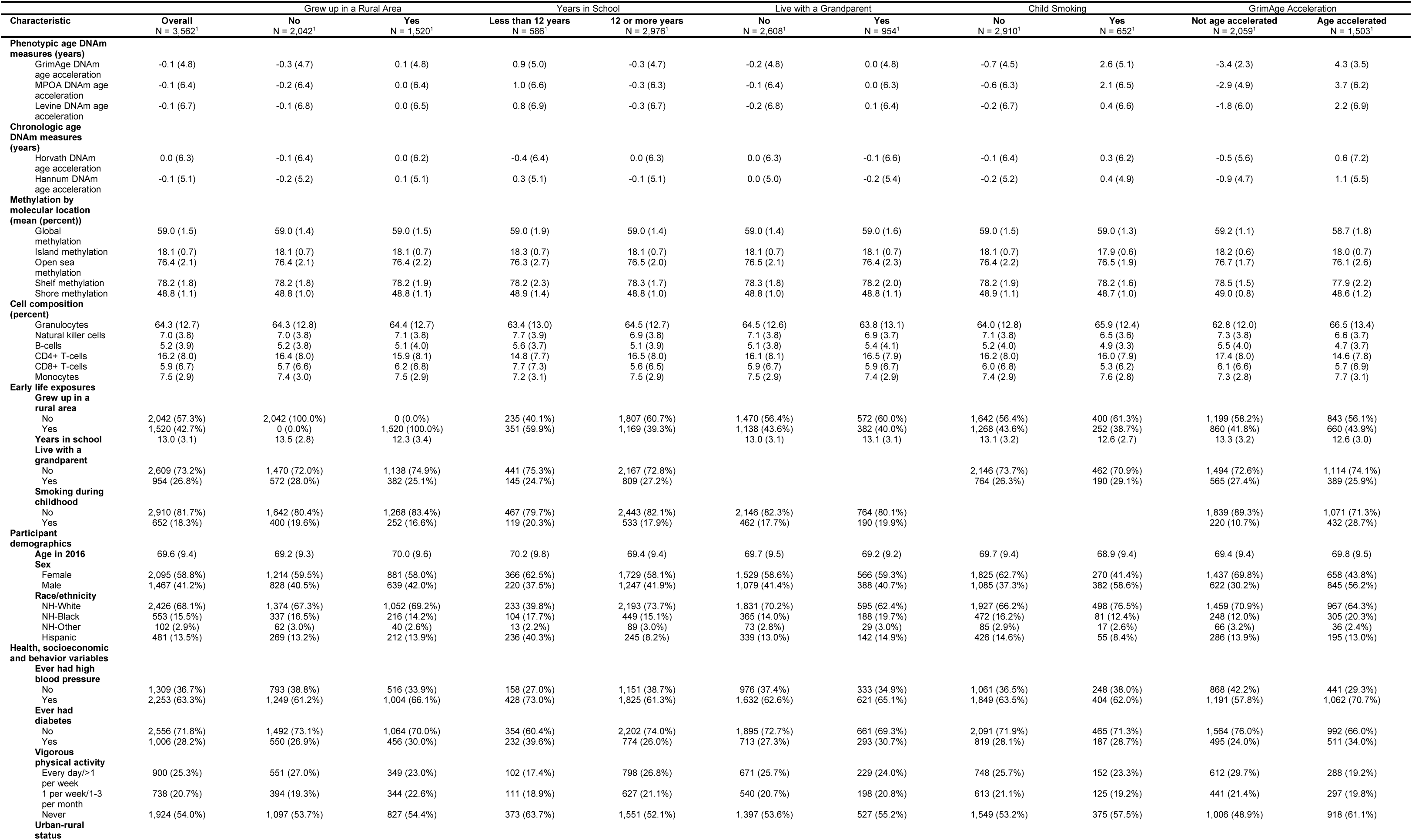
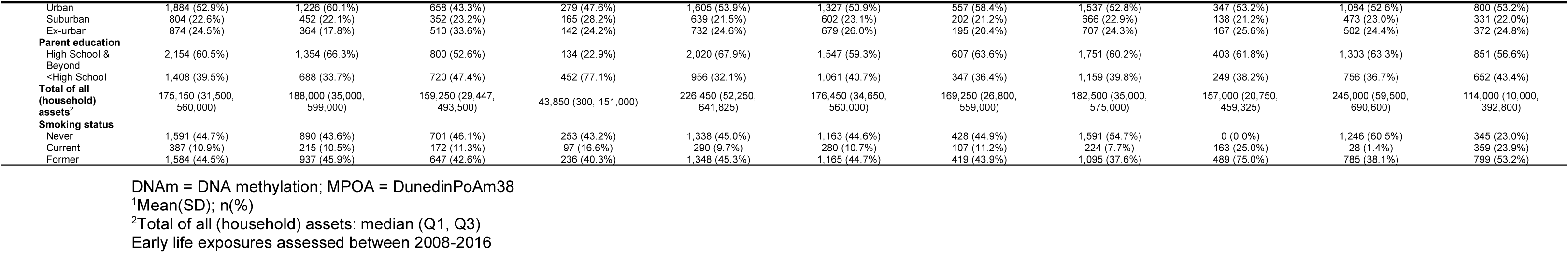
Bivariate descriptive statistics of participants from the U.S. Health and Retirement Study with early life exposures and GrimAge DNA methylation age acceleration among our analytic sample (N=3,562)

The DNA methylation age acceleration measures were positively and significantly correlated with each other (Range: r = 0.09 to r = 0.63) (**Supplemental Figure 2)**. We mostly saw weak correlations between our categorical variables (**Supplemental Figure 3**). Participants with less than 12 years of education and those who smoked during childhood both had higher GrimAge (**Figure 2**). Approximately 42.2% of our analytic sample had accelerated GrimAge, and these participants were more likely to report growing up in a rural area, spending fewer years in school, not living with a grandparent, and smoking during childhood, compared to those without accelerated GrimAge (**Table 1**). The mean percent global methylation at each molecular location was similar across exposures (**Table 1**).

**Figure 2.**
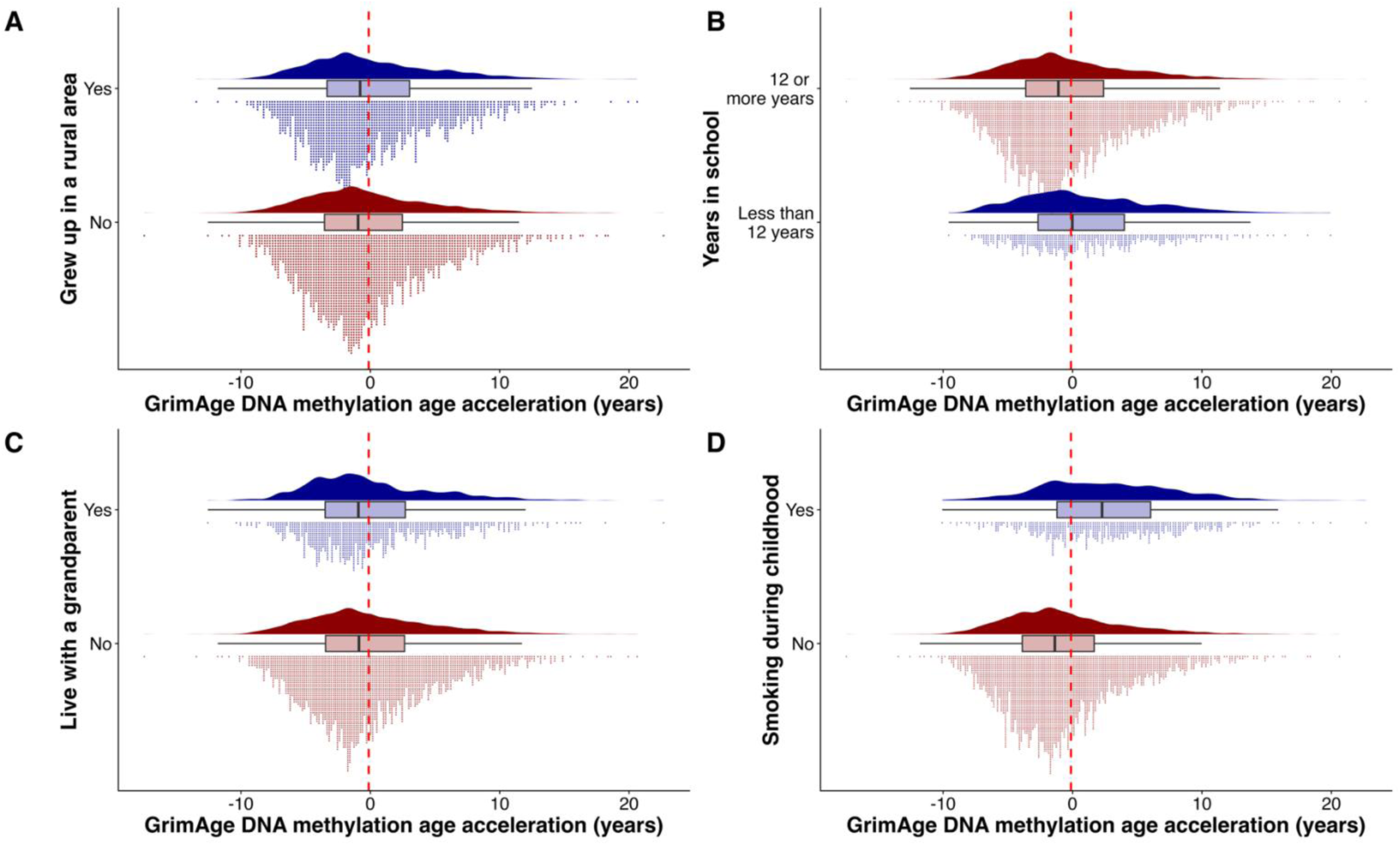
Raincloud plot of participants from the U.S. Health and Retirement Study with early life exposures and available GrimAge DNA Methylation Acceleration from the 2016 Venous Blood Study (N=3,562) Early life exposures were assessed between 2008-2016. Panel A shows the distribution of responses for whether a participant reported growing up in a rural area or not among our analytic sample. Panel B shows the distribution of responses for whether a participant had 12 or more years in school or less than 12 years among our analytic sample. Panel C shows the distribution of responses for whether a participant reported living with a grandparent or not among our analytic sample. Panel D shows the distribution of responses for whether a participant reported smoking during childhood or not among our analytic sample. Based on the above figure, we see no difference between response groups for variables growing up in a rural area and whether a participant lived with a grandparent during childhood. Conversely, those who reported having 12 or more years of schooling had lower GrimAge acceleration compared to those who reported spending less than 12 years in school. Also, we see that those who reported smoking during childhood had higher GrimAge acceleration compared to those who reported not smoking during childhood. This aligns with the bivariate associations we found above between childhood smoking and GrimAge acceleration.

### Early life exposures and DNA methylation age acceleration

We ran models to test associations between each of our early life exposures and each DNA methylation age acceleration measure. For our unadjusted models, growing up in a rural area, years in school, and smoking during childhood were each significantly associated with GrimAge acceleration (**Table 2**). Additionally, years in school and smoking during childhood were each significantly associated with MPOA age acceleration. Years in school was individually associated with Levine age acceleration, while smoking during childhood was individually associated with Hannum age acceleration. Living with a grandparent was not significantly associated with any of the age acceleration measures.

**Table 2.** Linear regression models assessing the association between four early life exposures and five DNA methylation age clocks in the Health and Retirement Study (N=3,562)

|  | GrimAge acceleration | MPOA acceleration | Levine age acceleration | Horvath age acceleration | Hannum age acceleration |
| --- | --- | --- | --- | --- | --- |
|  | beta [95%CI] FDR | beta [ 95%CI] FDR | beta [95%CI] FDR | beta [95%CI] FDR | beta [95%CI] FDR |
| **Grew up in a rural area** |  |  |  |  |  |
| Unadjusted | **0.36 [0.04, 0.68] 0.026** | 0.17 [-0.26, 0.59] 0.44 | 0.11 [-0.34, 0.55] 0.64 | 0.08 [-0.34, 0.50] 0.72 | 0.29 [-0.05, 0.63] 0.11 |
| Socioeconomic and behavior | 0.17 [-0.06, 0.41] 0.31 | 0.08 [-0.25, 0.41] 0.73 | -0.05 [-0.48, 0.37] 0.92 | -0.06 [-0.45, 0.32] 0.88 | 0.13 [-0.18, 0.45] 0.78 |
| **Years in school** |  |  |  |  |  |
| Unadjusted | **0.19 [0.15, 0.24] 3.65x10<sup>-14</sup>** | **0.21 [0.14, 0.27] 2.20x10<sup>-9</sup>** | **0.12 [0.05, 0.19] 0.0023** | -0.06 [-0.12, 0.01] 0.20 | 0.05 [-0.003, 0.10] 0.069 |
| Socioeconomic and behavior | **0.10 [0.06, 0.14] 2.53 x 10<sup>-5</sup>** | **0.11 [0.05, 0.17] 2.3x10<sup>-3</sup>** | 0.04 [-0.04, 0.12] 0.75 | -0.07 [-0.14, -0.004] 0.19 | 0.0006 [-0.06, 0.06] 0.99 |
| **Live with a grandparent** |  |  |  |  |  |
| Unadjusted | 0.13 [-0.22, 0.49] 0.47 | 0.16 [-0.31, 0.63] 0.50 | 0.26 [-0.24, 0.75] 0.31 | -0.10 [-0.58, 0.37] 0.92 | -0.22 [-0.60, 0.16] 0.53 |
| Socioeconomic and behavior | 0.06 [-0.20, 0.31] 0.87 | 0.02 [-0.34, 0.38] 0.97 | 0.23 [-0.23, 0.70] 0.75 | 0.01 [-0.41, 0.43] 0.99 | -0.03 [-0.38, 0.31] 0.97 |
| **Smoking during childhood** |  |  |  |  |  |
| Unadjusted | **3.34 [2.95, 3.73] 1.08x10<sup>-60</sup>** | **2.64 [2.10, 3.17] 1.40x10<sup>-21</sup>** | 0.64 [0.07, 1.21] 0.054 | 0.41 [-0.13, 0.95] 0.28 | **0.54 [0.11, 0.98] 0.030** |
| Socioeconomic and behavior | **0.65 [0.32, 0.97] 4.8x10<sup>-4</sup>** | 0.28 [-0.18, 0.74] 0.38 | -0.15 [-0.74, 0.44] 0.83 | -0.06 [-0.60, 0.47] 0.90 | -0.14 [-0.58, 0.30] 0.83 |

FDR = False discovery rate; multiple testing correction for the 20 tests performed (4 exposures and 5 outcomes)
Socioeconomic and behavior models include age, sex, race/ethnicity, health variables (ever had high blood pressure, ever had diabetes, vigorous physical activity), socioeconomic and behavior variables (Urban-rural status, Parent Education, Total of all [household] assets, Adult smoking status), cell composition (Granulocytes, Natural Killer cells, B-cells, CD4+ T-cells, and CD8+ T-cells), and sample plate
Early life exposures assessed between 2008-2016
Bolded text denotes a statistically significant finding (FDR adjusted p-value<0.05)

Comparing our demographic and health models, we saw a mostly consistent magnitude and direction of association between each early life exposure and age acceleration measure (**Supplemental Table 4)**. Years in school was no longer significantly associated with Levine age acceleration after adjusting for health covariates. Further, smoking during childhood was no longer significantly associated with Hannum age acceleration after adjusting for demographic model covariates.

In our socioeconomic and behavior models, years in school was still significantly associated with GrimAge and MPOA age acceleration (**Table 2 and Supplemental Figure 4**). A one year decrease in the years spent in school was significantly associated with 0.10 years higher GrimAge (95% CI: 0.06, 0.14) and 0.11 years higher MPOA (95% CI: 0.05, 0.17) DNA methylation age acceleration. Smoking during childhood was still significantly associated with GrimAge acceleration, but not MPOA age acceleration. Smoking during childhood was associated with 0.65 years higher GrimAge DNA methylation age acceleration (95% CI: 0.32, 0.97). Additionally, we observed a significant attenuation in the magnitude of the estimates for smoking during childhood, which is likely due to the adjustment for adult smoking status. No significant associations were observed for growing up in a rural area or living with a grandparent among the 5 DNA methylation age acceleration measures. However, we observed a change in the estimates direction of association between growing up in a rural area and Levine age acceleration after adjusting for adult socioeconomic and behavior variables. We also observed this for living with a grandparent and GrimAge and MPOA age acceleration.

### Early life exposures and global DNA methylation

Next, we evaluated associations between each early life exposure with global DNA methylation calculated in five molecular locations (Global, Island, Open Sea, Shelf, and Shore). Across our models, living with a grandparent had lower percent methylation at each molecular location (**Supplemental Figure 4 and Supplemental Table 3**). Results from our socioeconomic and behavior adjusted models were null across all five molecular locations.

### Early life exposures and site-specific DNA methylation

Slight inflation was observed for growing up in a rural area, with a lambda value of 1.15, and years in school, with a lambda value of 1.11 (**Supplemental Figure 5**). No inflation was observed for living with a grandparent, with a lambda value of 0.87, and smoking during childhood, with a lambda value of 0.85. For years spent in school, 49689 sites had DNA methylation difference (p-value<0.05), and among these sites, 53.5% had lower DNA methylation (**Figure 3**). No sites reached epigenome-wide significance for any of the three other early life exposures. Full model output for each exposure from our socioeconomic and behavior EWAS models are available in **Supplemental Table 7**. We found years spent in school significantly associated (adjusted p-value<0.05) with 231 DNA methylation sites (**Supplemental Table 7**). The top five sites from our socioeconomic and behavior EWAS for each exposure are presented in **Table 3**. The second top CpG site for years in school was cg07318158 (0.47 percent lower methylation; FDR p-value: 1.69 × 10^-9^), which annotated to the gene *OTUD7B* (**Table 3**). Results from our health and demographic adjusted models are available in the supplemental materials (**Supplemental Table 8 and Supplemental Table 9**). We observed 49 nominally associated (p<1×10^-4^) sites for growing up in a rural area, 23 nominal sites for living with a grandparent, and 20 nominal sites for smoking during childhood. Among our nominally associated (p<1×10^-4^) sites, we found no overlap between each of our early life exposures from our socioeconomic and behavior EWAS models (**Supplemental Figure 6**). Additionally, we observed weak correlations between exposures (**Supplemental Figure 9**).

**Figure 3.**
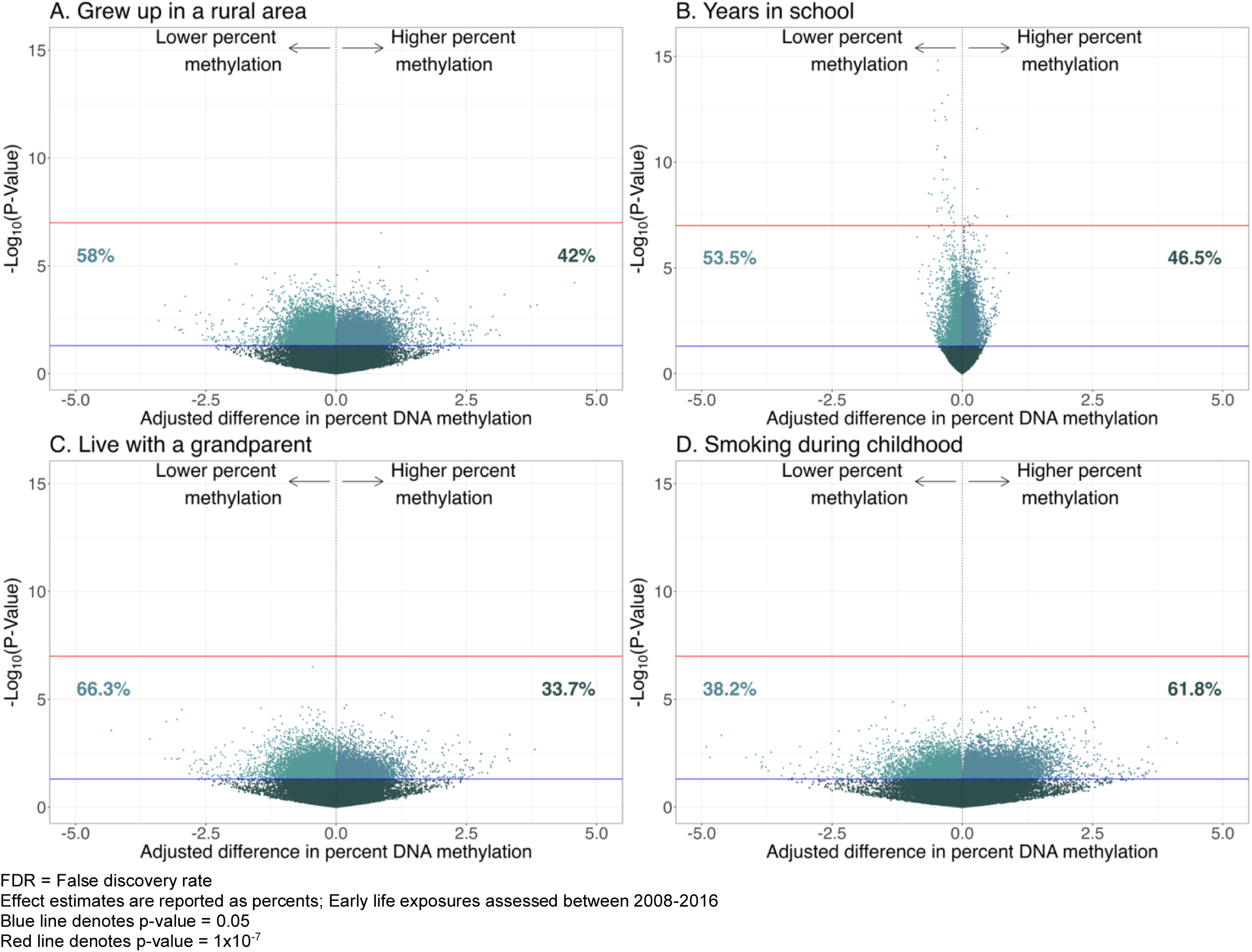
Single CpG site association P-values from our socioeconomic and behavior models for four early life exposures and later life DNA methylation in the Health and Retirement Study

**Table 3.**
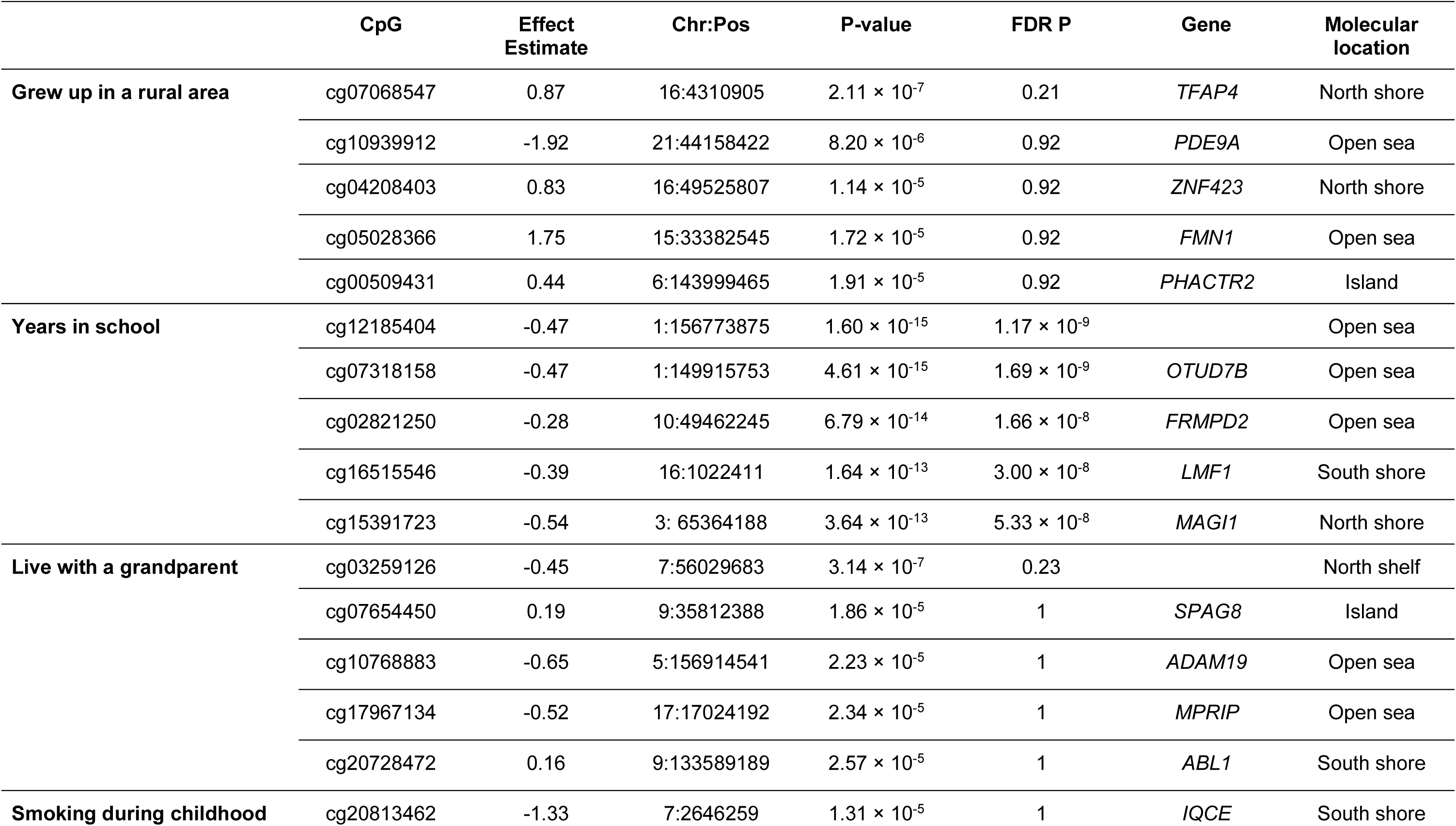
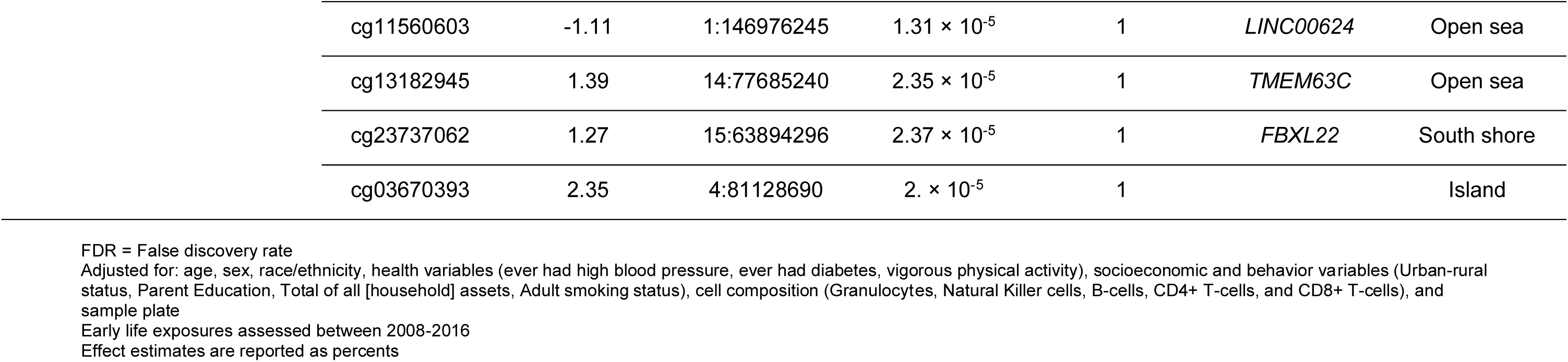
Top 5 significant CpG sites (p-value < 10^-4^) from the epigenome-wide association study showing the difference in percent methylation for each early life exposure on later life DNA methylation in the Health and Retirement Study.

To understand the impact of multiple exposures on DNA methylation, we ran our socioeconomic and behavior adjusted models, including all exposures in one mixtures model (**Supplemental Table 12**). We observed similar model fit statistics and covariate inflation between our mixtures DNA methylation age acceleration and global methylation models, and our independent socioeconomic and behavior early life exposure models. The proportion of the variance in the outcome explained (R^2^) was consistent across the models (**Supplemental Table 6**), and we observed adjusted VIF values less than 5, suggesting low multicollinearity of covariates. Additionally, our mixtures and individual socioeconomic and behavior models were consistent in magnitude. Similar to our individual early life EWAS models adjusted for socioeconomic and behavior covariates, we found no overlapping sites (p<1×10^-4^) across each early life exposure in our mixture models (**Supplemental Figure 7**). We found a total of 650 unique sites (p<1×10^-4^) in the mixtures early life exposure site-specific models that were not shared across our individual early life exposures, with 21 unique sites for growing up in a rural area, 576 for years in school, 23 for living with a grandparent, and 30 for smoking during childhood.

### Replication and additional analyses

To understand whether there are shared DNA methylation sites across two of our early life exposures and two prior EWAS,^22,25^ we examined overlap of DNA methylation sites with exposure-specific p-values < 1×10^-4^ (**Supplemental Figure 8**). We found no associated (p<1×10^-4^) sites overlap between our early life exposure years in school and the prior EWAS on maternal educational attainment (**Supplemental Figure 8**).^22^ For our early life exposure of smoking during childhood and the prior EWAS on former and current smoking^25^, we found an overlap at cg13481974 for former smoking and at cg25636159 for current smoking. Both sites annotated to the gene *MYO1G*. Further, we compared effect estimates and unadjusted p-values from all tested sites for the socioeconomic and behavior models of our years in school and smoking during childhood exposures to the same two prior EWAS (**Supplemental Figure 10 and Supplemental Table 10**). We observed a moderate correlation (r = -0.5) between effect estimates from all tested sites for the socioeconomic and behavior adjusted EWAS models of years in school and the prior EWAS on maternal education (**Supplemental Figure 10**). We observed a moderate correlation for former smoking (r = 0.42) and a weak correlation for current smoking (r = 0.17) with our early life exposure of smoking during childhood (**Supplemental Figure 10**). This suggests that among the sites we compared, smoking during childhood in our study is moderately correlated with former smoking behaviors and weakly correlated with current adult smoking behaviors in the prior EWAS.

All sites identified from our socioeconomic and behavior EWAS model were utilized to examine potential enrichment of gene ontology pathways (**Table 4 and Supplemental Table 11**). Sites identified for growing up in a rural area were significantly enriched in pathways associated with cellular membrane processing, sorting, and vesicle transport of proteins and lipids, including during endocytosis (**Table 4)**. Pathways identified for years in school were involved in cellular changes due to metal ion stimuli, processes involved in embryonic development, and the organization of extracellular structures (**Table 4)**. For living with a grandparent, pathways identified were involved in the breakdown of proteins, regulation of hormones within and between cells, and ATP processes (**Table 4)**. Finally, smoking during childhood was significantly enriched for pathways involved in exocytosis, regulation of nervous system development, and processes involved in cell adhesion and structure (**Table 4)**. However, we found these top five pathways to be unique for each early life exposure.

**Table 4.**
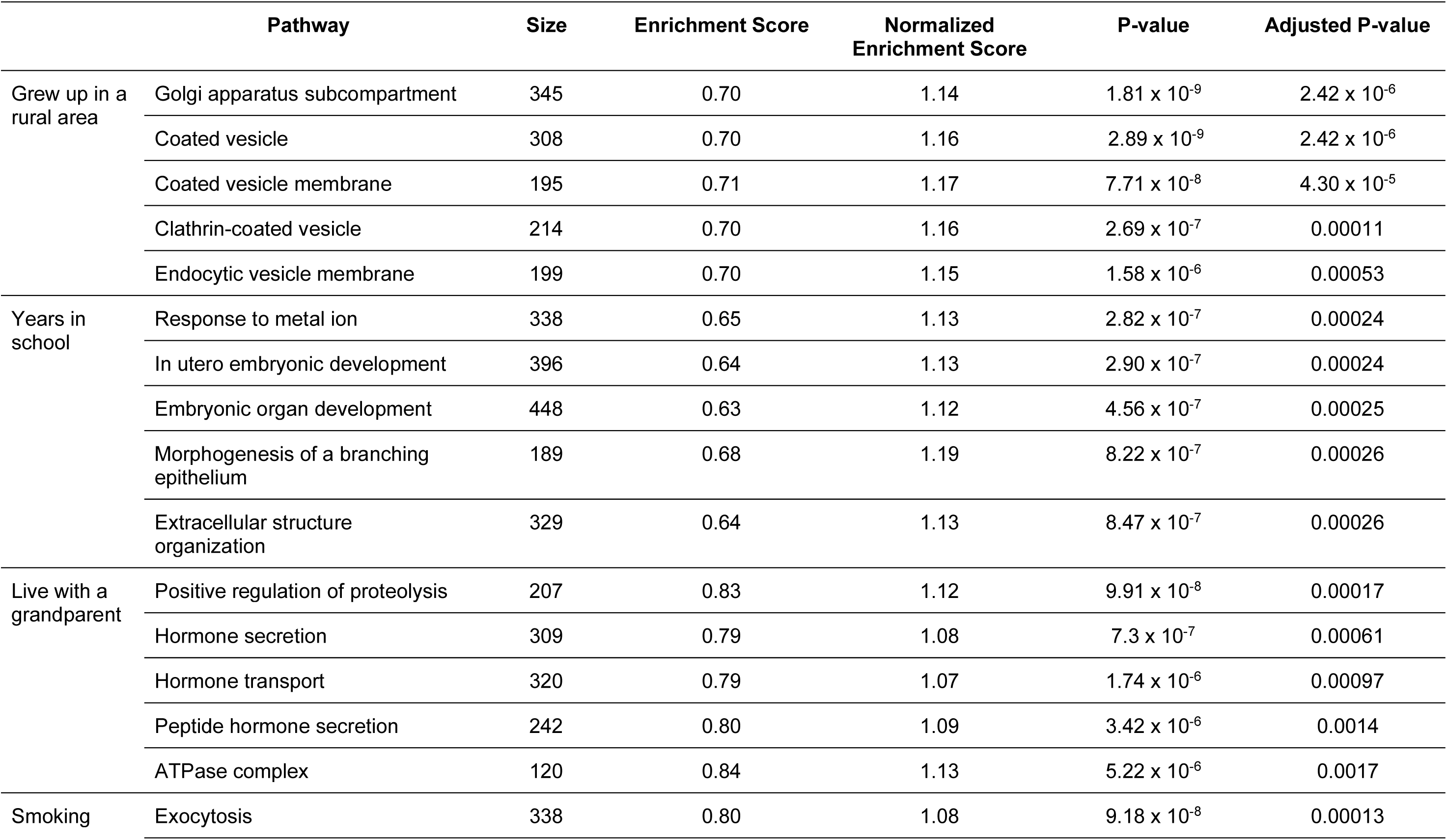
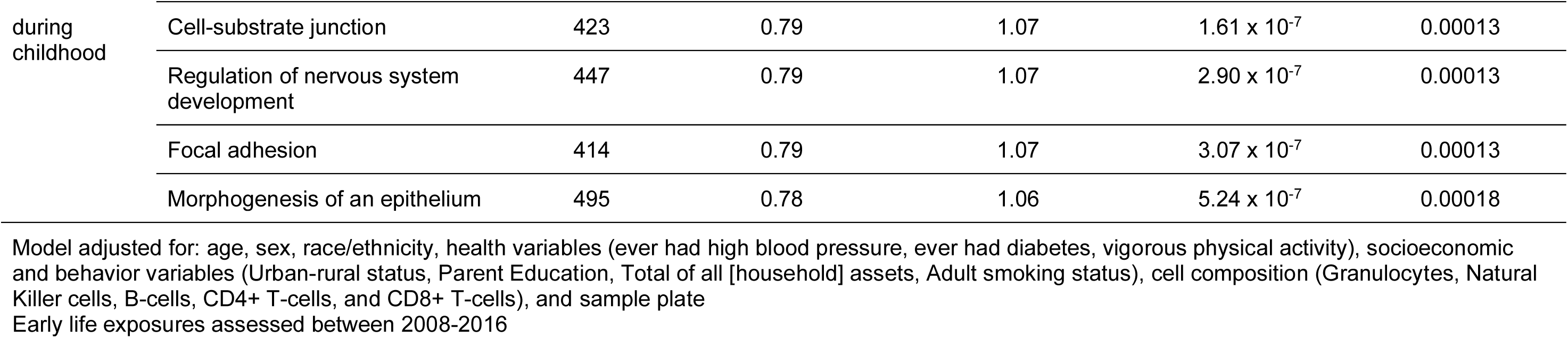
Top 5 Gene Ontology pathway enrichment analysis results from our socioeconomic and behavior models for four early life exposures and later life DNA methylation in the Health and Retirement Study.

## Discussion

Among a cohort of middle-aged and older U.S. adults (N=3,562), we sought to understand the impact of four early life exposures on later life DNA methylation age acceleration, global and site-specific methylation, and underlying biological processes. We hypothesized that growing up in a rural area, fewer years in school, living with a grandparent, and smoking during childhood would be significantly associated with differences in DNA methylation in later life, and adjusting for adult health, socioeconomic, and behavior variables would attenuate these associations. Fewer years spent in school and smoking during childhood were associated with accelerated GrimAge DNA methylation age after adjusting for demographic, health, socioeconomic, and behavior variables. In our EWAS analysis, fewer years spent in school was significantly associated with 231 CpG sites (adjusted p-value<0.05). Results for our other early life exposures were not statistically significant. Findings from our mixtures early life exposure model results were consistent in magnitude with our individual early life exposure models across our three DNA methylation measures. For our replication analysis, we found overlap (p<1×10^-4^) with 2 CpG sites for our smoking during childhood exposure with a prior meta-analysis on adult cigarette smoking behavior,^25^ but no overlap (p<1×10^-4^) between our years in school exposure and a prior meta-analysis on maternal educational attainment.^22^ Our gene set enrichment analysis identified five significantly enriched pathways unique for each exposure. We hypothesized that these early life experiences can co-occur in individuals and subsequently impact later life DNA methylation compared to when we examine the individual level effects of these exposures. Our results suggest that each early life exposure has unique DNA methylation signatures that do not overlap.

Years spent in school was consistently and significantly associated with GrimAge and MPOA age acceleration. Our findings align with a previous study among a sample of adults from the HRS and the Multi-Ethnic Study of Atherosclerosis (MESA).^20^ Results suggest that fewer years of education in both cohorts is associated with more GrimAge and faster MPOA age acceleration. Although we did not find associations between years in school and Levine age acceleration, fewer years of education in their analysis of the HRS was significantly associated with more Levine age acceleration. Another study conducted in the HRS found a similar association between personal years of education and DNA methylation aging.^19^ Higher years of education were significantly associated with less GrimAge and slower MPOA age acceleration. Additionally, higher parental years of education were independently associated with less Horvath and GrimAge DNA methylation age acceleration, as personal years of education increased. The direction of the associations in these two prior studies is consistent with our study findings for GrimAge, MPOA, and Levine. However, in a cohort of older adults in Mexico with past early life socioeconomic and environmental adversities, individuals were younger chronologically relative to our sample of participants, and more years of schooling were associated with less Horvath and Levine age acceleration but no difference in their Hannum age compared to those with fewer years of schooling.^60^ This differs slightly in direction from our findings, where we reported that fewer years in school was associated with more Levine and less Horvath age acceleration. Differences in the direction of association across prior studies and our study could be due to the adjustment of adult health covariates in our socioeconomic and behavior adjusted model compared to those in our study.

Our estimate of the association between smoking during childhood and GrimAge acceleration was consistent in direction and magnitude with a prior study in the HRS examining smoking in youth and GrimAge acceleration.^21^ Another study found associations between early life smoking and faster MPOA and more Levine age acceleration.^61^ The magnitude and direction of association for the MPOA and Levine DNA methylation age acceleration measures were consistent with our socioeconomic and behavior model results. In another study among MESA participants, early life adversity, specifically parental deprivation, was associated with GrimAge accelerated DNA methylation age, and smoking behaviors mediated a portion of this association.^62^

We did not find any significant associations between any of our early life exposures and global methylation. This suggests that our early life exposures are impacting DNA methylation at specific sites rather than broad differences in DNA methylation. This highlights the importance of future studies examining early life exposures to examine multiple measures of DNA methylation to capture different patterns across the genome.

In a prior EWAS examining years in school, nominal associations were found for high (≥ 9 years) vs. low (< 9 years) years of schooling.^60^ Another study conducted only in MESA found a significant negative association between maternal education and DNA methylation in five stress and inflammatory genes.^63^ In an EWAS meta-analysis of educational attainment across 27 European ancestry cohorts, ranging between mean ages of 26.6 to 79.1 years, findings reveal that 9 sites were significantly associated with educational attainment.^64^ However, these results were attenuated upon stratifying by smoking status. Further, the authors find that the results overall are biased due to the correlation of educational attainment with other confounding variables (e.g., smoking). This is consistent with our EWAS results for years in school and smoking during childhood. Model coefficients between our health adjusted model and our socioeconomic and behavior model were significantly attenuated due to the adjustment of adult smoking.

In an Australian cohort of 17 year olds, self-reported adolescent smoking among those not exposed to maternal smoking was not associated with any tested sites after adjusting for age, sex, maternal age, cell counts, birthweight, gestational weight gain, maternal alcohol consumption during pregnancy, maternal school level, maternal pre-pregnancy BMI, family income during pregnancy, and batch.^23^ This is consistent with our findings for self-reported smoking during childhood and later life site-specific methylation. However, in the meta-analyses examining current and former smoking compared to never smoking, there were significantly associated sites after adjusting for sex, age, cell type proportions, and cohort.^25^

Growing up in a rural area and living with a grandparent were not significantly associated with any of our DNA methylation measures in our socioeconomic and behavior models. Both of these exposures are underexplored in the literature for their association with DNA methylation, but have been found to impact later life cognitive function and co-occur with other adverse early life experiences.^65–67^ Research should continue to explore these early life exposures and other social factors in large, diverse cohorts for their potential to impact later life site-specific methylation.

In our replication analysis for smoking during childhood, we tested for replication with a prior study on adult smoking status.^25^ We found significant overlap between previously identified sites for former and current smoking, compared to never smoking, with our sites for smoking during childhood. Among these, cg13481974 in former smokers and cg25636159 in current smokers annotated to *MYO1G*. Prior studies on this gene found consistent associations with early life and adult smoking, including persistent effects from childhood to late adolescence.^24,68^ Additional studies are needed to investigate our findings in each early life exposure and older populations.

Top sites identified in our EWAS were unique for each of our early life exposures. Sites for growing up in a rural area were significantly enriched for biological processes involved in cellular membrane processes and vesicle transport. Years in school was enriched for pathways involved in metal ion stimuli and embryonic development. Our early life exposure, living with a grandparent, was enriched for pathways involved in the regulation of protein breakdown and ATP processing. Smoking during childhood had pathways involved in exocytosis and nervous system development.

### Limitations

Interpretation of our study’s findings should also take into consideration some limitations. A key limitation is that DNA methylation was only measured at a single point in time, and we only considered adult factors as precision variables. The literature suggests that early life experiences impact later life health and that adult health, socioeconomic, and behavior factors influence these associations independently and jointly. These factors have been found to attenuate the association between early life experiences and later life DNA methylation, potentially due to their ability to act as mediators, confounders, and/or precision variables.^26,62,61^ Future studies should collect early and midlife exposures along with DNA methylation at multiple time points across the life course to better capture the impact of these experiences on later life DNA methylation patterns and subsequent health outcomes. Additionally, examination of adult factors beyond precision covariates can help disentangle the complex associations between early life experiences and DNA methylation, including the implementation of identified sites as biomarkers for potential subsequent adverse health outcomes. This would allow for an assessment of the temporality and alterations of DNA methylation levels at different periods across the life course and the impact of adult factors along the causal path to more deeply understand the independent and combined impact of early and midlife experiences on later life DNA methylation and health. Although we framed our early life exposures as worse, it is possible that this generation of adults had different social and behavioral norms (e.g., smoking during childhood) and interpretations of what these early life exposures mean relative to our current assessment.^69–71^ Future studies should consider the framing and examination of early life factors as not inherently harmful, depending on the age of their population. Our excluded sample of participants was found to be older chronologically, have higher mean GrimAge and MPOA acceleration, be more likely to report growing up in a rural area, have fewer personal years in school, have parents with lower educational attainment, and have lower median household assets in adulthood compared to our included sample. Due to these differences among the included and excluded samples, it is possible that our results are underestimating the true association between early life experiences and DNA methylation due to the exclusion of an older and more vulnerable group. Finally, our exposures were assessed retrospectively, which introduces the potential for bias. To account for this, we checked for inconsistencies between the early life exposure of smoking and the adult behavior of smoking. We found participants who reported smoking during childhood but never smoking in adulthood and removed them to reduce the potential for information bias. Future studies should consider this in the design and implementation of their study, including potentially prospectively collecting early life information. This could potentially identify sensitive periods and proximal risk factors for differences in DNA methylation during early development that can be intervened upon to prevent later life consequences.

### Strengths

This study has several strengths that contribute to current gaps in the literature. We had a demographically diverse sample of participants relative to prior research studies. Our study is one of the few that have explored early life experiences with later life site-specific methylation. Further, we reported a large, robust analytic sample (N=3,562) of participants who passed all DNA methylation QC and preprocessing steps, allowing for the reduction in potential spurious findings due to residual technical artifacts. This study adds to the current literature on early life exposures and later life DNA methylation by implementing multiple measures of DNA methylation. Prior studies have focused on single measures of DNA methylation, which may limit the ability to fully understand the impact of these exposures on DNA methylation patterns across the genome.^20–22,25,61,64^ Further, we adjusted for demographic, adult health, socioeconomic, and behavior covariates in our models. This allowed us to account for potential biased findings while adjusting for these variables, which have been suggested to affect the association between early life exposures and DNA methylation as potential confounders, mediators, and precision variables. Although we observed moderate correlations between years in school and parent education, along with smoking during childhood and adult smoking behaviors, we still saw significant associations with GrimAge acceleration. Although our findings are consistent with prior studies that examined associations between broad self-reported early life exposures, like early life socioeconomic status, early life smoking, parental abuse/neglect, parental divorce, childhood residential moves, and childhood trouble with the police, with measures of DNA methylation in midlife and older adulthood,^20,21,72–75^ our study examined underexplored early life exposures beyond parental abuse and neglect and socioeconomic status. In our comparisons of effect estimates from our socioeconomic and behavior EWAS models, we found weak correlations between exposures. We observed distinct signatures for each early life experience in our mixtures model, suggesting that differences in DNA methylation sites may vary by exposure. Thus, it is important to evaluate these separately for potential unique biological pathways that may mediate associations with later life health outcomes. Further, studies can use these unique signatures to create exposure-specific DNA methylation biomarkers.

## Conclusion

Overall, our findings suggest that each early life exposure has distinct DNA methylation patterns that are important to understand separately for their potential impact on later life health. DNA methylation clocks are responsive to social and environmental factors, and accelerated biological aging is suggested to be associated with an increased risk of morbidity and mortality. The association between early life exposures and later life differences in DNA methylation may be influenced by midlife behavioral and socioeconomic factors. Studies find that early life exposure to stressors can increase the risk of poor adult health behaviors (e.g., diet and/or smoking), which directly affects stress response regulatory systems that lead to subsequent physiological changes through the activation of genes.^76–78^ Prior studies have highlighted the implications of early life experiences on later life morbidity and mortality, including an increased risk of decreased cognitive function in later life.^76,79–81^ Various mechanisms, like gene expression and inflammation, have been explored to understand the impact of early life experiences on later life health.^63,77,82^ Future research should continue to explore factors relating to individual social, behavioral, and environmental exposures across the life course to better understand potential vulnerable periods for intervention and mitigation of later life health risks and premature mortality.

## Acknowledgments

We want to thank the staff and participants of the Health and Retirement Study.

## Data availability statement

Datasets used for the analysis, including the DNA methylation age measures, are available from the Health and Retirement Study website (https://hrs.isr.umich.edu/data-products). Access to the Health and Retirement Study DNA methylation data measured on the Infinium Methylation EPIC (v1.0) BeadChip array requires an application to the National Institute on Aging Genetics of Alzheimer’s Disease Data Storage Site (NIAGADS) for use (Accession Number: NG00153.v1). All code files used in this manuscript are available on a GitHub repository (https://github.com/bakulskilab/EarlyLife_DNAm_HRS).

