## Supplemental Figures and Tables for "Early life experiences are associated with later life DNA methylation signatures in the Health and Retirement Study"

---

### Supplemental Figures and Tables

**Supplemental Figure 1.** Scatter plot of participants from the U.S. Health and Retirement Study chronologic age in 2016 and GrimAge DNA methylation age acceleration

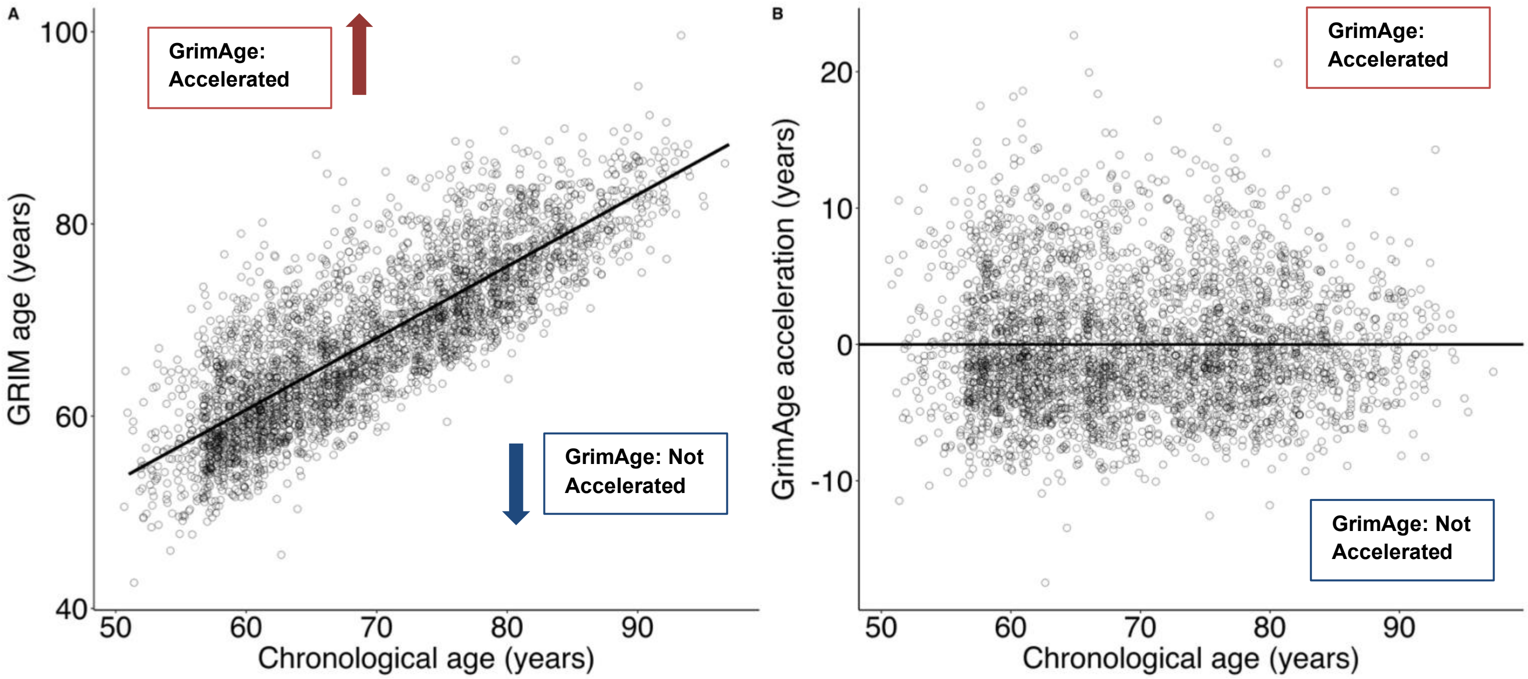


The scatterplot on the left (A) depicts the calculation of the GrimAge residuals (also known as GrimAge DNAm (DNA methylation) age acceleration) by regressing chronological age onto GrimAge. Positive residuals are represented by the points above the line of best fit reflecting accelerated DNAm GrimAge. Not accelerated DNAm GrimAge represents points below the line. We used GrimAge residuals as the outcome in our DNAm age acceleration analysis. On the right (B) is a scatterplot of chronological age and the GrimAge residuals we calculated in the left plot.

**Supplemental Table 1.** Description of early life exposure variables questionnaires and DNA methylation clocks information from the 2016 Venous Blood Study of participants from the U.S. Health and Retirement Study

| Variable label | HRS variable name | Description[1-3] |
|---|---|---|
| Grew up in a rural area | childrural | Were you living in a rural area most of the time when you were~about age 10?” |
| Years in school | SCHLYRS | From 0-17, what is the number of years you were in school? |
| Live with a Grandparent | LIVEGPAR | Did you ever live in the same household with a grandparent for a year or more before age 17? |
| Smoking during childhood | CHSMOKE | Did you regularly smoke cigarettes while you were in grade school or high school? By "regularly" we mean at least one cigarette a day for most days of the week, for six months or more. |
| GrimAge DNAm age and age acceleration (residuals) | DNAMGRIMAGE | GrimAge was developed by Lu et al. (2019) and is based on 7 DNAm surrogate markers of plasma proteins and smoking pack-years. This study utilized an older adult sample from the Framingham Heart Study and was validated in 3 additional cohorts. Peripheral blood samples were processed and measured on the Illumina 450k array, and the model was trained on both the 450k and EPIC arrays.[4] |
| MPOA DNAm age and age acceleration (residuals) | MPOA | Developed by Belsky et al. (2020), the methylation Pace of Aging (MPOA), also known as DunedinPoAm38, is a score based on 38 CpG sites in whole blood. The study included 954 middle-aged adult participants from three waves of the Dunedin Study, with additional validation cohorts. The model was trained on the Illumina 450k array.[5] |
| Levine DNAm age and age acceleration (residuals) | LEVINE_DNAMAGE | Developed by Levine et al. (2018) using clinical data from adults over 20 years old who participated in the NHANES III as the training sample. Participants 20 and older from NHANES IV and 5 other cohorts served as validation samples. This clock is based on 513 CpGs from various tissues and cell types, including whole blood, and was trained on the Illumina EPIC array.[6] |
| Horvath DNAm age and age acceleration (residuals) | HORVATH_DNAMAGE | Developed by Horvath (2013), this clock is based on data from 8,000 samples across age groups from 82 Illumina DNAm array datasets. It incorporated 353 CpGs from 51 tissue and cell types, including peripheral and whole blood. The model was trained on the Illumina 27k and 450k arrays.[7] |

| | | |
|---|---|---|
| Hannum DNAm age and age acceleration (residuals) | HANNUM_DNAMAGE | The Hannum clock, developed by Hannum et al. (2013), is based on 71 CpGs from a single tissue type in whole blood. The study utilized a sample of 656 adults ranging from young to older age from two cohorts. The model was trained on the Illumina 450k array.[8] |
| **DNAm:** DNA methylation; **HRS**: Health and Retirement Study; **MPOA**: Methylation pace of aging<br>Early life exposures assessed between 2008-2016 | | |

**Supplemental Table 2.** Descriptive statistics for the included and excluded sample of participants from the U.S. Health and Retirement Study with early life exposures and available DNA methylation data from the 2016 Venous Blood Study

| **Characteristic** | **Overall**<br>N = 4,018[1] | **Excluded**<br>N = 456[1] | **Included**<br>N = 3,562[1] | **p-value**[2] |
|---|---|---|---|---|
| **Phenotypic age DNAm measures (years)** | | | | |
| GrimAge DNAm age acceleration | 0.0 (4.8) | 1.0 (4.5) | -0.1 (4.8) | <0.001 |
| MPOA DNAm age acceleration | 0.0 (6.4) | 0.7 (6.7) | -0.1 (6.4) | 0.020 |
| Levine DNAm age acceleration | 0.0 (6.8) | 0.7 (7.8) | -0.1 (6.7) | 0.077 |
| **Chronologic age DNAm measures (years)** | | | | |
| Horvath DNAm age acceleration | 0.0 (6.4) | 0.3 (7.1) | 0.0 (6.3) | >0.9 |
| Hannum DNAm age acceleration | 0.0 (5.2) | 0.4 (6.1) | -0.1 (5.1) | 0.4 |
| **Methylation by molecular location (mean (percent))** | | | | |
| Global methylation | 59.0 (1.5) | 59.0 (1.4) | 59.0 (1.5) | 0.2 |
| Island methylation | 18.1 (0.7) | 18.2 (0.7) | 18.1 (0.7) | 0.2 |
| Open sea methylation | 76.4 (2.1) | 76.4 (2.0) | 76.4 (2.1) | 0.061 |
| Shelf methylation | 78.2 (1.8) | 78.2 (1.7) | 78.2 (1.8) | 0.020 |
| Shore methylation | 48.8 (1.1) | 48.8 (1.1) | 48.8 (1.1) | 0.7 |
| Missing | 38 | 38 | 0 | |
| **Cell composition (percent)** | | | | |
| Granulocytes | 64.3 (13.0) | 63.6 (15.2) | 64.3 (12.7) | 0.8 |
| Natural killer cells | 7.1 (3.9) | 7.5 (4.6) | 7.0 (3.8) | 0.14 |
| B-cells | 5.2 (4.3) | 5.9 (6.9) | 5.2 (3.9) | 0.086 |
| CD4+ T-cells | 16.0 (8.0) | 14.6 (8.0) | 16.2 (8.0) | <0.001 |
| CD8+ T-cells | 6.0 (6.8) | 6.7 (7.5) | 5.9 (6.7) | 0.039 |
| Monocytes | 7.5 (2.9) | 7.4 (3.2) | 7.5 (2.9) | 0.3 |
| **Early life exposures** | | | | |
| **Grew up in a rural area** | | | | <0.001 |
| Yes | 1,717 (43.6%) | 197 (52.8%) | 1,520 (42.7%) | |
| No | 2,218 (56.4%) | 176 (47.2%) | 2,042 (57.3%) | |
| Missing | 83 | 83 | 0 | |
| **Years in school** | 12.8 (3.2) | 11.3 (3.5) | 13.0 (3.1) | <0.001 |
| Missing | 19 | 19 | 0 | |
| **Live with a grandparent** | | | | 0.024 |
| Yes | 1,096 (27.4%) | 142 (32.0%) | 954 (26.8%) | |
| No | 2,910 (72.6%) | 302 (68.0%) | 2,608 (73.2%) | |
| Missing | 12 | 12 | 0 | |

| Characteristic | Overall<br>N = 4,018[1] | Excluded<br>N = 456[1] | Included<br>N = 3,562[1] | p-value[2] |
|---|---|---|---|---|
| **Smoking during childhood** | | | | 0.017 |
| Yes | 748 (18.8%) | 96 (23.2%) | 652 (18.3%) | |
| No | 3,227 (81.2%) | 317 (76.8%) | 2,910 (81.7%) | |
| Missing | 43 | 43 | 0 | |
| **Participant demographics** | | | | |
| **Age in 2016** | 69.8 (9.7) | 72.0 (11.3) | 69.6 (9.4) | <0.001 |
| **Sex** | | | | 0.2 |
| Male | 1,669 (41.5%) | 202 (44.3%) | 1,467 (41.2%) | |
| Female | 2,349 (58.5%) | 254 (55.7%) | 2,095 (58.8%) | |
| **Race/ethnicity** | | | | <0.001 |
| NH-White | 2,669 (66.5%) | 243 (53.6%) | 2,426 (68.1%) | |
| NH-Black | 657 (16.4%) | 104 (23.0%) | 553 (15.5%) | |
| NH-Other | 122 (3.0%) | 20 (4.4%) | 102 (2.9%) | |
| Hispanic | 567 (14.1%) | 86 (19.0%) | 481 (13.5%) | |
| Missing | 3 | 3 | 0 | |
| **Health, socioeconomic, and behavior variables** | | | | |
| **Ever had high blood pressure** | | | | 0.013 |
| Yes | 2,568 (63.9%) | 315 (69.2%) | 2,253 (63.3%) | |
| No | 1,449 (36.1%) | 140 (30.8%) | 1,309 (36.7%) | |
| Missing | 1 | 1 | 0 | |
| **Ever had diabetes** | | | | 0.013 |
| Yes | 1,160 (28.9%) | 154 (33.9%) | 1,006 (28.2%) | |
| No | 2,856 (71.1%) | 300 (66.1%) | 2,556 (71.8%) | |
| Missing | 2 | 2 | 0 | |
| **Vigorous physical activity** | | | | 0.10 |
| Every day/>1 per week | 1,001 (24.9%) | 101 (22.1%) | 900 (25.3%) | |
| 1 per week/1-3 per month | 822 (20.5%) | 84 (18.4%) | 738 (20.7%) | |
| Never | 2,195 (54.6%) | 271 (59.4%) | 1,924 (54.0%) | |
| **Urban-rural status** | | | | 0.10 |
| Urban | 2,109 (52.5%) | 225 (49.6%) | 1,884 (52.9%) | |
| Suburban | 927 (23.1%) | 123 (27.1%) | 804 (22.6%) | |
| Ex-urban | 980 (24.4%) | 106 (23.3%) | 874 (24.5%) | |
| Missing | 2 | 2 | 0 | |
| **Parent education** | | | | <0.001 |
| High School & Beyond | 2,239 (59.8%) | 85 (45.9%) | 2,154 (60.5%) | |

| Characteristic | Overall<br>N = 4,018[1] | Excluded<br>N = 456[1] | Included<br>N = 3,562[1] | p-value[2] |
|---|---|---|---|---|
| <High School | 1,508 (40.2%) | 100 (54.1%) | 1,408 (39.5%) | |
| Missing | 271 | 271 | 0 | |
| **Total of all (household) assets**[2] | 161,100 (25,100, 526,000) | 79,700 (4,000, 287,000) | 175,150 (31,500, 560,000) | <0.001 |
| **Smoking status** | | | | 0.009 |
| Current | 455 (11.4%) | 68 (15.7%) | 387 (10.9%) | |
| Former | 1,776 (44.5%) | 192 (44.3%) | 1,584 (44.5%) | |
| Never | 1,764 (44.2%) | 173 (40.0%) | 1,591 (44.7%) | |
| Missing | 23 | 23 | 0 | |

[1]Mean(SD); n(%)

[2]Total of all (household) assets: median (Q1,Q3)

[3]Wilcoxon rank sum test; Fisher's exact test

Early life exposures assessed between 2008-2016

**Supplemental Figure 2.** Correlation matrix of Pearson correlation coefficients for the continuous variables from the analysis of early life exposures and later life DNA Methylation in the Health and Retirement Study

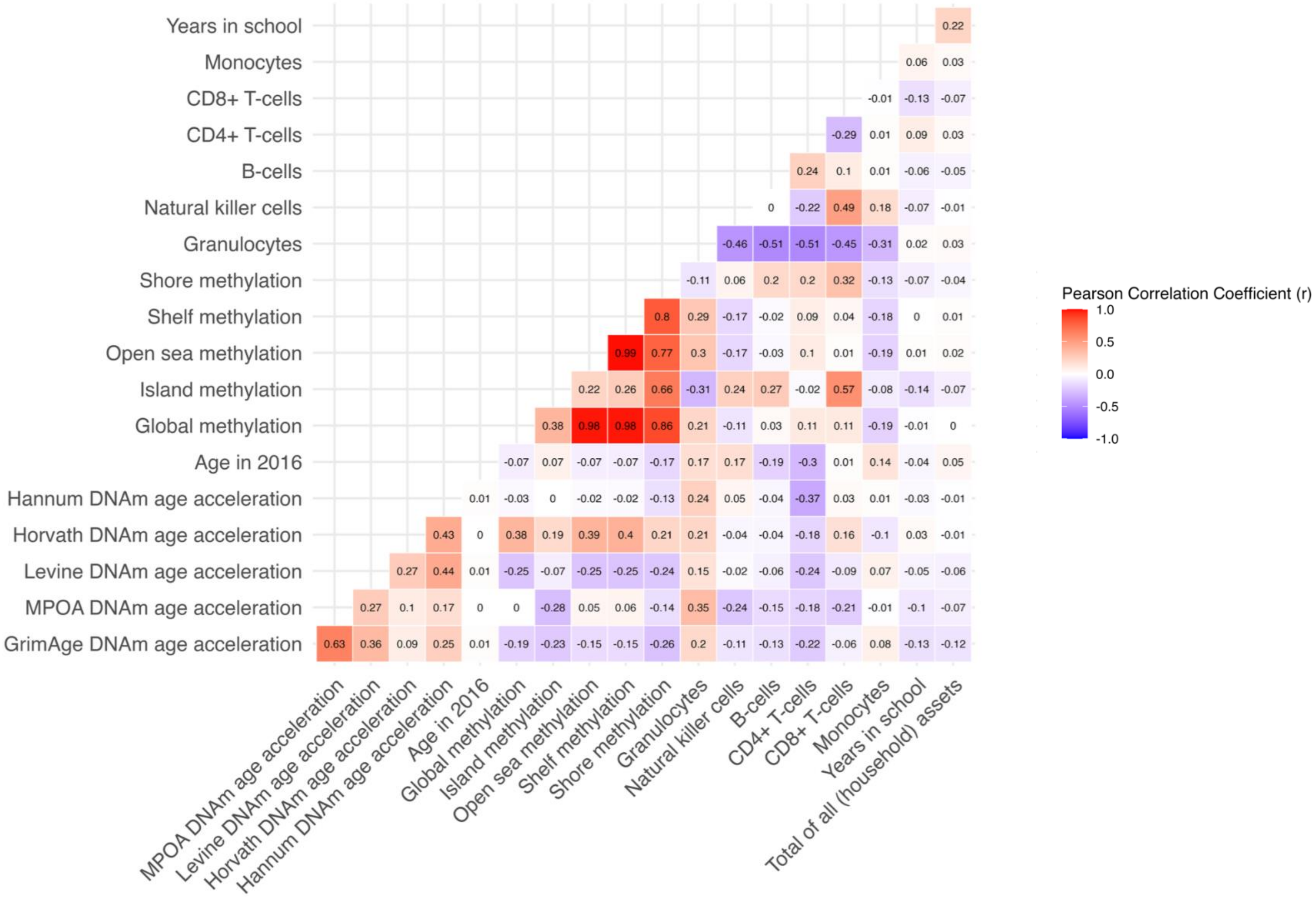


MPOA = DunedinPoAm38

**Supplemental Figure 3.** Correlation matrix of Cramer's V coefficients for the categorical variables from the analysis of early life exposures and later life DNA Methylation in the Health and Retirement Study

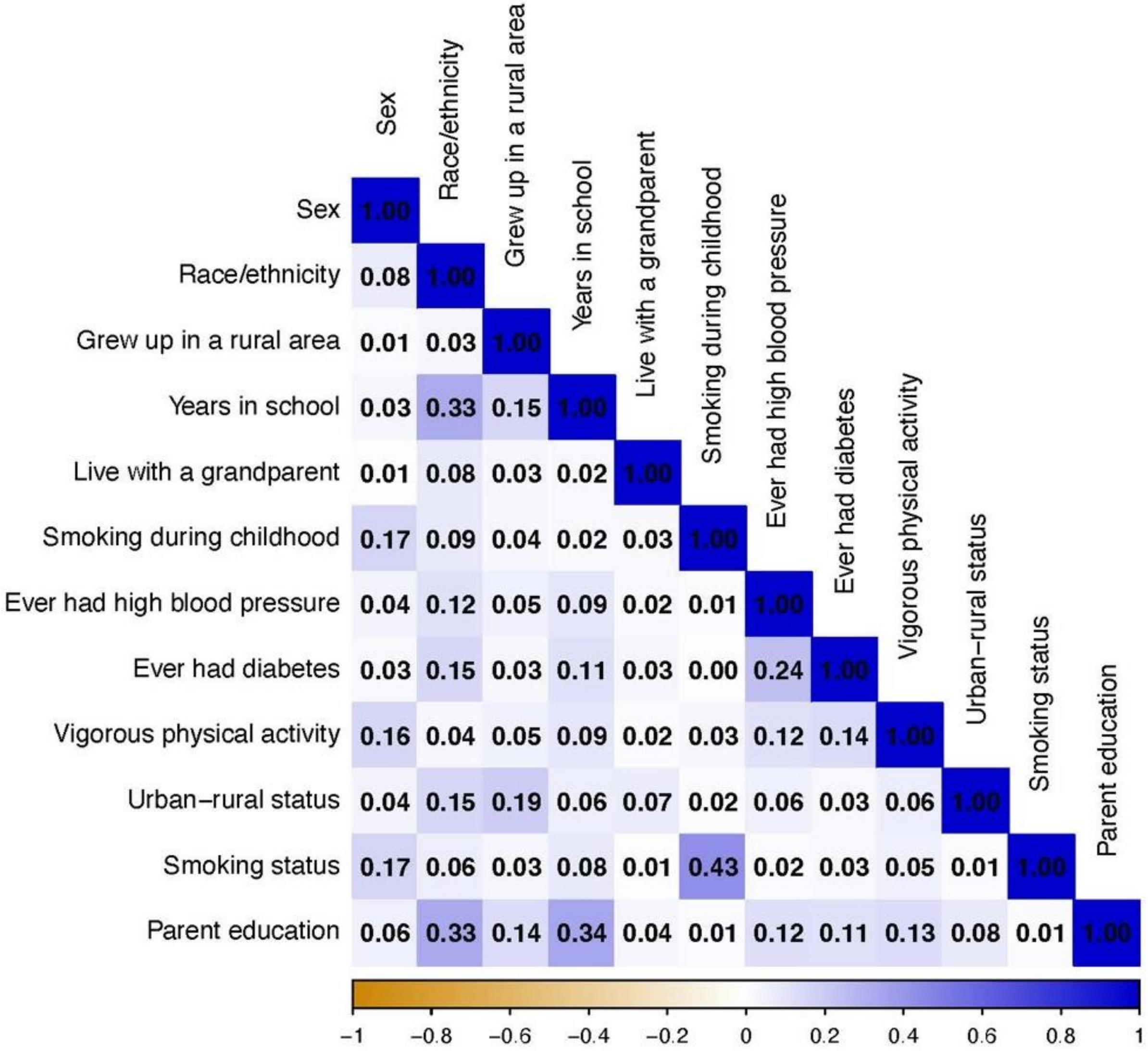

**Supplemental Figure 4.** Forest plots of socioeconomic and behavior linear regression models assessing the association between four early life exposures with DNA methylation age acceleration and mean global methylation measures by molecular location in the Health and Retirement Study

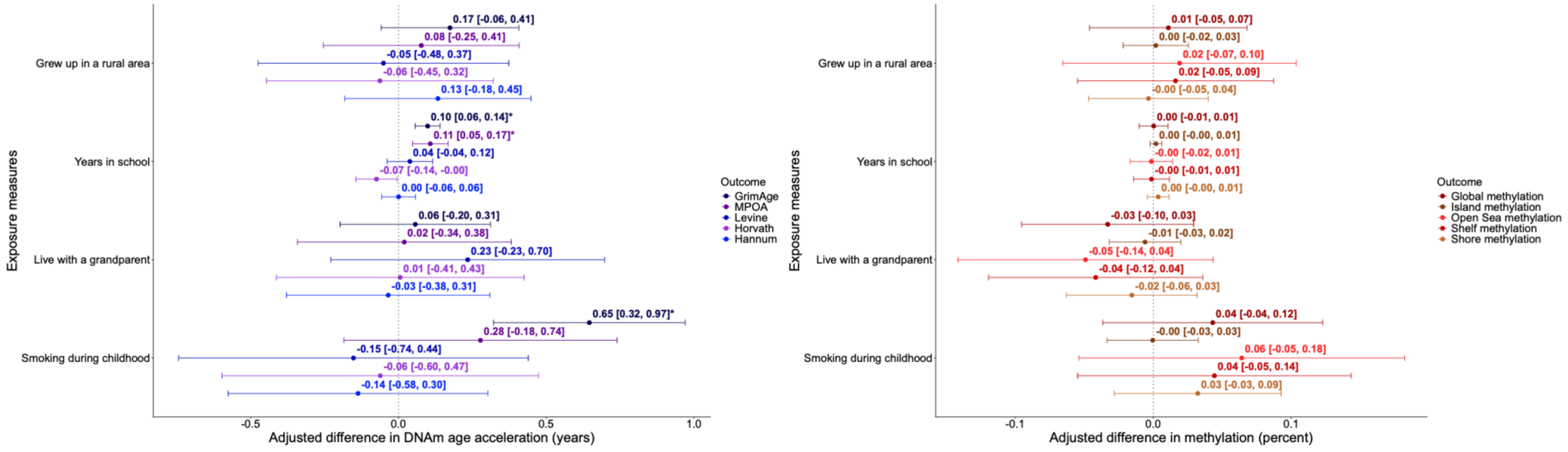


Early life exposures assessed between 2008-2016

**Supplemental Table 3**. Results from linear regression models assessing the association between four early life exposures and global methylation measures by molecular location in the Health and Retirement Study (N=3,562)

| | Mean global methylation | Mean island methylation | Mean open sea methylation | Mean shelf methylation | Mean shore methylation |
|---|---|---|---|---|---|
| | **beta [95%CI] FDR** | **beta [95%CI] FDR** | **beta [ 95%CI] FDR** | **beta [95%CI] FDR** | **beta [95%CI] FDR** |
| **Grew up in a rural area** | | | | | |
| Unadjusted | -0.01 [-0.11, 0.09] 0.87 | 0.03 [-0.02, 0.07] 0.22 | -0.02 [-0.16, 0.12] 0.79 | -0.01 [-0.13, 0.11] 0.91 | -0.02 [-0.09, 0.05] 0.59 |
| Socioeconomic and behavior | 0.01 [-0.05, 0.07] 0.84 | 0.002 [-0.02, 0.03] 0.90 | 0.02 [-0.07, 0.10] 0.80 | 0.02 [-0.05, 0.09] 0.87 | -0.003 [-0.05, 0.04] 0.89 |
| **Years in school** | | | | | |
| Unadjusted | 0.01 [-0. 01, 0.02] 0.39 | **0.03 [0.02, 0.04] $1.23\times10^{-16}$** | -0.01 [-0.03, 0.02] 0.54 | -0.003 [-0.02, 0.02] 0.80 | **0.02 [0.01, 0.03] $9.83\times10^{-5}$** |
| Socioeconomic and behavior | 0.00036 [-0.01, 0.01] 0.96 | 0.002 [-0.002, 0.006] 0.44 | -0.001 [-0.02, 0.01] 0.94 | -0.001 [-0.01, 0.01] 0.96 | 0.003 [-0.004, 0.01] 0.51 |
| **Live with a grandparent** | | | | | |
| Unadjusted | -0.05 [-0.16, 0.06] 0.35 | -0.002 [-0.05, 0.05] 0.93 | -0.09 [-0.24, 0.07] 0.29 | -0.08 [-0.21, 0.06] 0.28 | -0.01 [-0.09, 0.07] 0.85 |
| Socioeconomic and behavior | -0.03 [-0.10, 0.03] 0.54 | -0.006 [-0.03, 0.02] 0.73 | -0.05 [-0.14, 0.04] 0.57 | -0.04 [-0.12, 0.04] 0.51 | -0.02 [-0.06, 0.03] 0.64 |
| **Smoking during childhood** | | | | | |
| Unadjusted | -0.056 [-0.18, 0.069] 0.38 | **-0.20 [-0.25, -0.14] $2.15\times10^{-11}$** | 0.03 [-0.15, 0.21] 0.73 | -0.002 [-0.16, 0.15] 0.98 | **-0.16 [-0.25, -0.07] 0.00045** |
| Socioeconomic and behavior | 0.043 [-0.036, 0.12] 0.52 | -0.0003 [-0.03, 0.03] 0.98 | 0.06 [-0.05, 0.18] 0.56 | 0.04 [-0.05, 0.14] 0.60 | 0.03 [-0.03, 0.09] 0.46 |

FDR = False discovery rate; multiple testing correction for the 20 tests performed (4 exposures and 5 outcomes)
Models adjusted for: age, sex, race/ethnicity, health variables (ever had high blood pressure, ever had diabetes, vigorous physical activity), socioeconomic and behavior variables (Urban-rural status, Parent Education, Total of all [household] assets, Adult smoking status), cell composition (Granulocytes, Natural Killer cells, B-cells, CD4+ T-cells, and CD8+ T-cells), and sample plate
Early life exposures assessed between 2008-2016
Bolded text denotes a statistically significant finding (FDR adjusted p-value <0.05)

**Supplemental Figure 5.** QQ plots assessing model diagnostics for our socioeconomic and behavior models of our four early life exposures and later life DNA methylation in the Health and Retirement Study

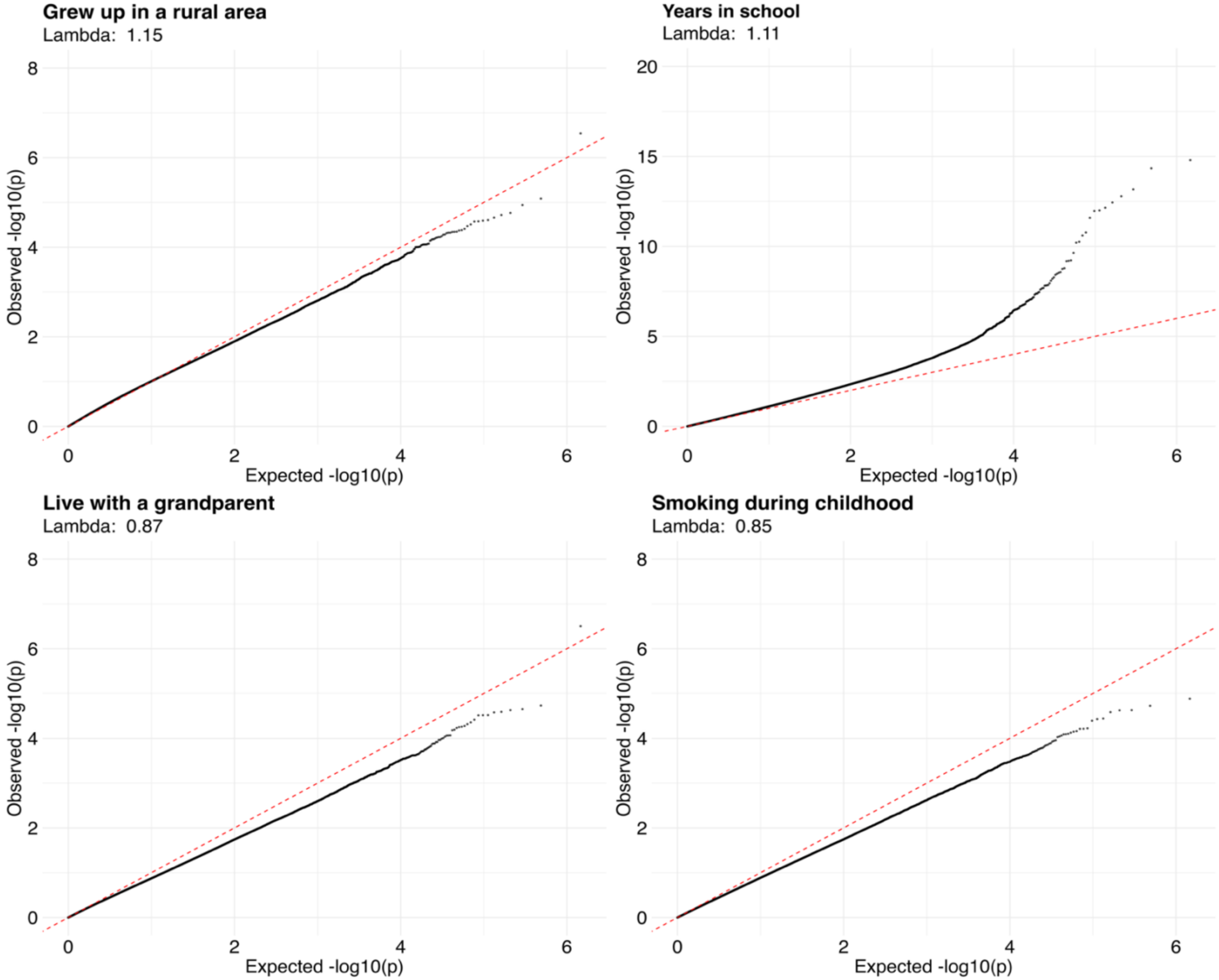


Lambda = Genomic inflation factor
Early life exposures assessed between 2008-2016

**Supplemental Table 4:** Linear regression models (Demographic and Health) assessing the association between four early life exposures and five DNA methylation age clocks in the Health and Retirement Study (N=3,562)

**File = Supplemental_Tables_4-5_AA_GM.xlsx**

**Supplemental Table 5:** Linear regression models (Demographic and Health) assessing the association between four early life exposures and five global methylation measures by molecular location in the Health and Retirement Study (N=3,562)

**File = Supplemental_Tables_4-5_AA_GM.xlsx**

**Supplemental Table 6:** R-squared values for individual early life exposure models and mixtures exposure model adjusted for socioeconomic and behavior variables assessing the association between four early life exposures and five DNA methylation age clocks in the Health and Retirement Study (N=3,562)

| | Grew up in rural area | Years in school | Live with a grandparent | Smoking during childhood | Mixtures |
|---|---|---|---|---|---|
| **GrimAge** | 0.508 | 0.511 | 0.508 | 0.510 | 0.513 |
| **MPOA** | 0.444 | 0.446 | 0.444 | 0.444 | 0.446 |
| **Levine** | 0.176 | 0.176 | 0.176 | 0.176 | 0.177 |
| **Horvath** | 0.247 | 0.248 | 0.247 | 0.247 | 0.248 |
| **Hannum** | 0.222 | 0.222 | 0.222 | 0.222 | 0.222 |

Adjusted for: age, sex, race/ethnicity, health variables (ever had high blood pressure, ever had diabetes, vigorous physical activity), socioeconomic and behavior variables (Urban-rural status, Parent Education, Total of all [household] assets, Adult smoking status), cell composition (Granulocytes, Natural Killer cells, B-cells, CD4+ T-cells, and CD8+ T-cells), and sample plate

**Supplemental Figure 6.** UpSet plot of top overlapping CpGs (p-value<$1 \times 10^{-4}$) for each of our early life exposures epigenome-wide association study model results adjusted for socioeconomic and behavior variables

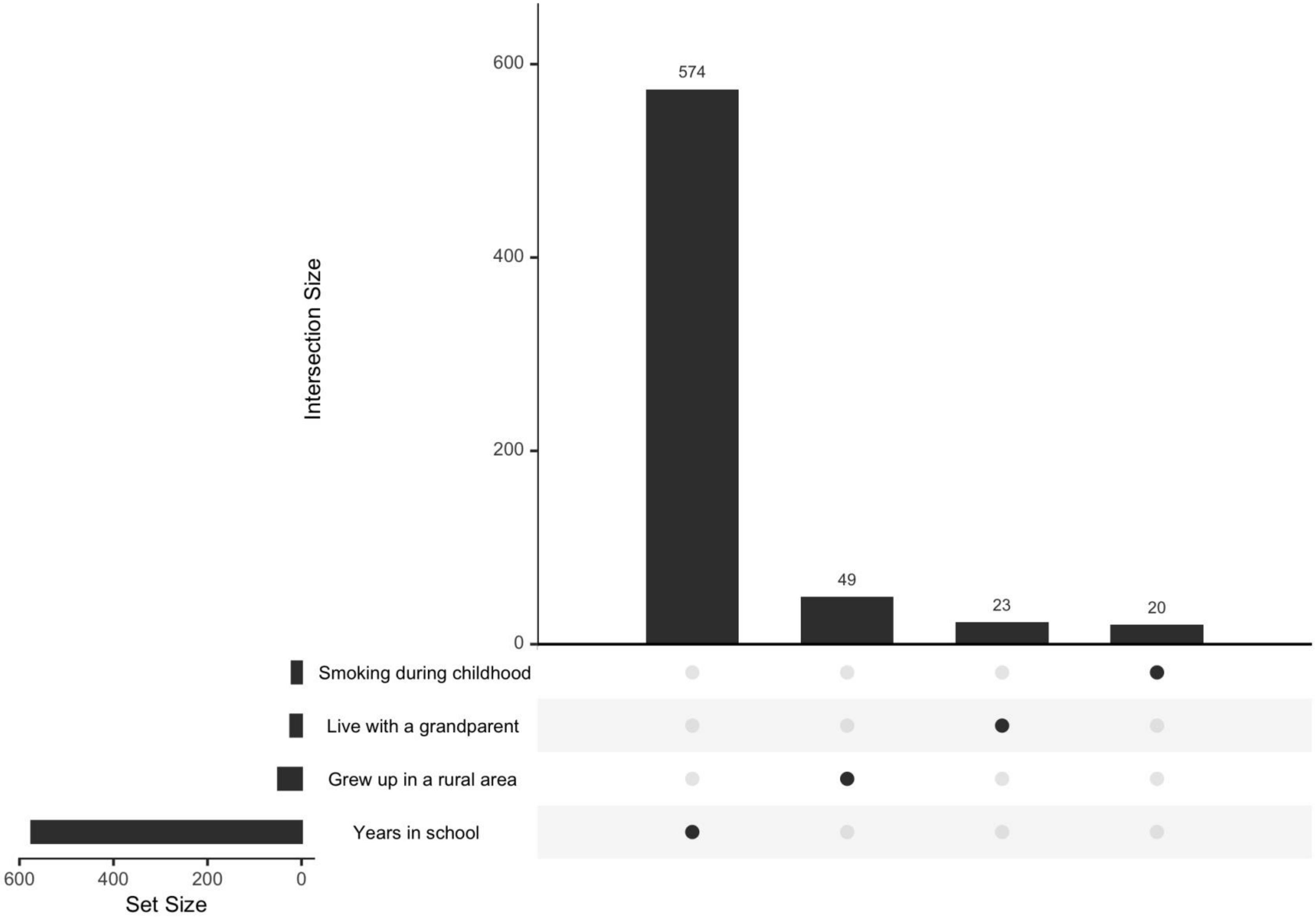

**Supplemental Figure 7.** UpSet plot of top overlapping CpGs (p-value<$1 \times 10^{-4}$) for our mixtures early life exposures epigenome-wide association study model results adjusted for socioeconomic and behavior variables

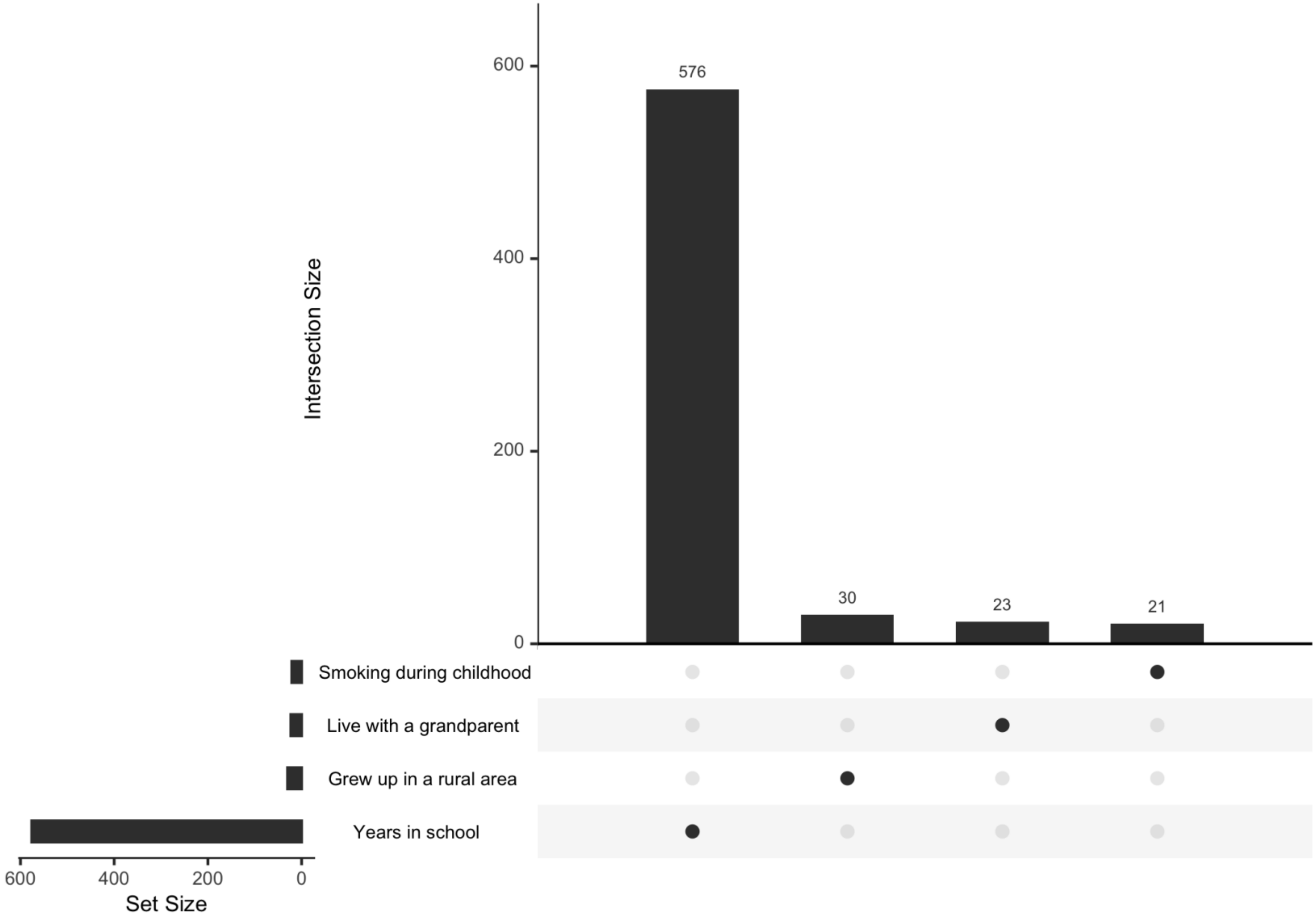

**Supplemental Figure 8.** UpSet plots of top overlapping CpGs (p-value<1x10$^{-4}$) for years in school and smoking during childhood epigenome-wide association study model results adjusted for socioeconomic and behavior variables with prior epigenome-wide association study results

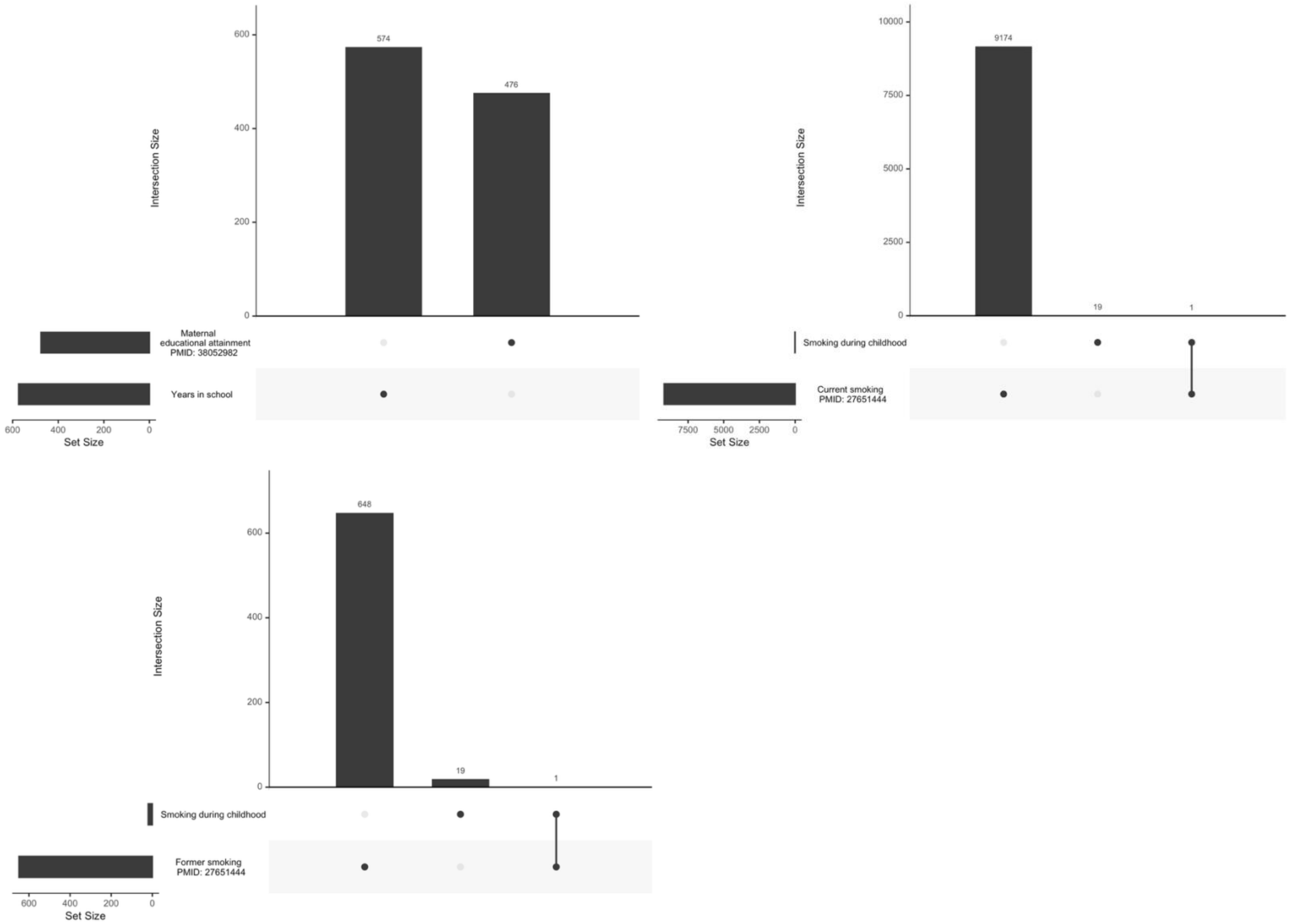

**Supplemental Figure 9.** Socioeconomic and behavior model effect estimate comparisons for all CpGs from an epigenome-wide association study of four early life exposures and later life DNA methylation in the Health and Retirement Study

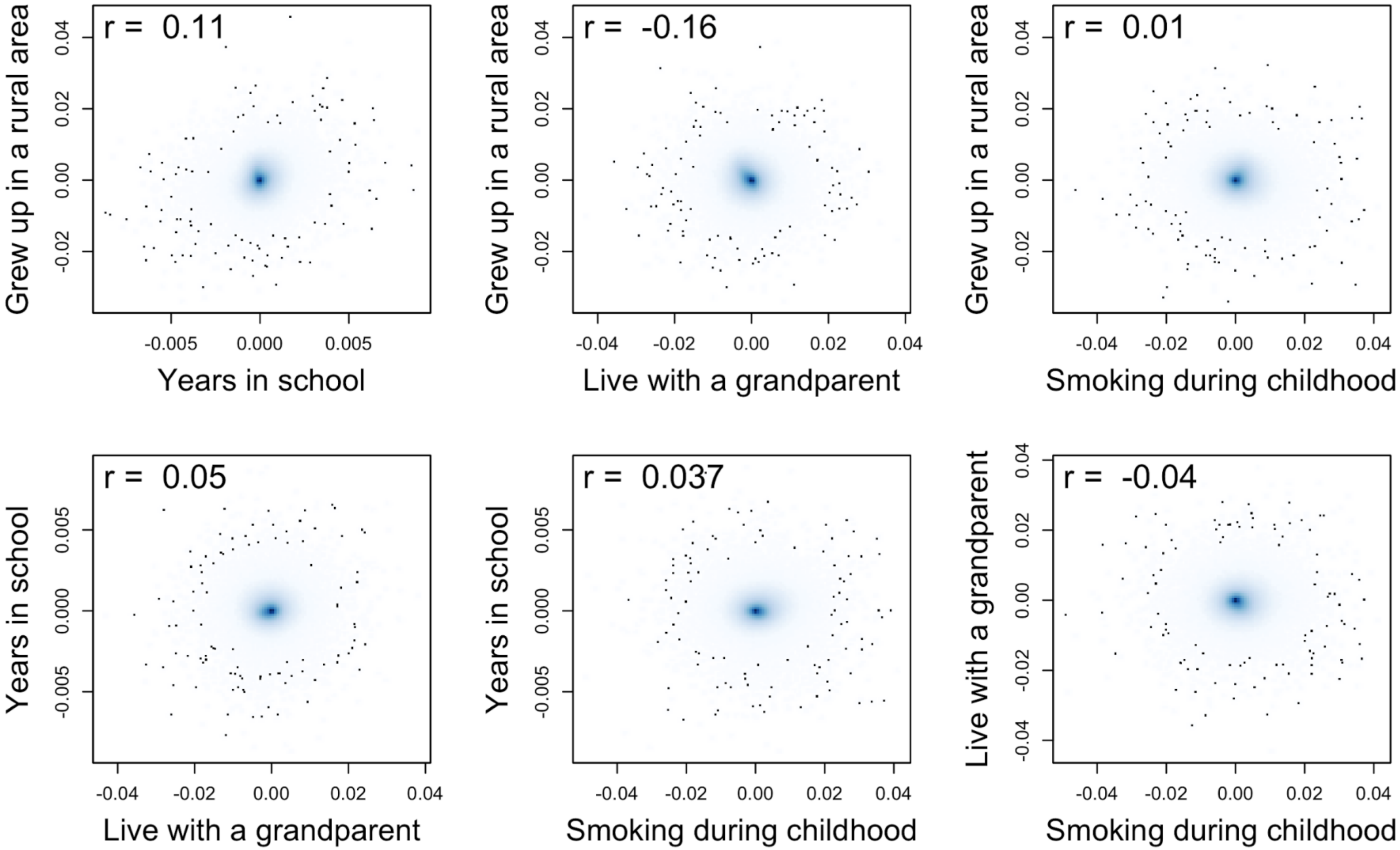


Early life exposures assessed between 2008-2016

**Supplemental Table 7.** Single sites from the socioeconomic and behavior epigenome-wide association study showing the difference in percent methylation for each early life exposure on later life DNA methylation in the Health and Retirement Study

**File = Supplemental Table 7- Single Site Results.xlsx**

**Supplemental Table 8.** Single sites from the health adjusted epigenome-wide association study showing the difference in percent methylation for each early life exposure on later life DNA methylation in the Health and Retirement Study

**File = Supplemental Table 8-Health Single Site Results.xlsx**

**Supplemental Table 9.** Single sites from the demographic adjusted epigenome-wide association study showing the difference in percent methylation for each early life exposure on later life DNA methylation in the Health and Retirement Study

**File = Supplemental Table 9-Demographic Single Site Results.xlsx**

**Supplemental Figure 10.** Socioeconomic and behavior model effect estimate comparisons for all CpGs from an epigenome-wide association study of years in school and smoking during childhood and later life DNA methylation in the Health and Retirement Study with prior epigenome-wide association study results

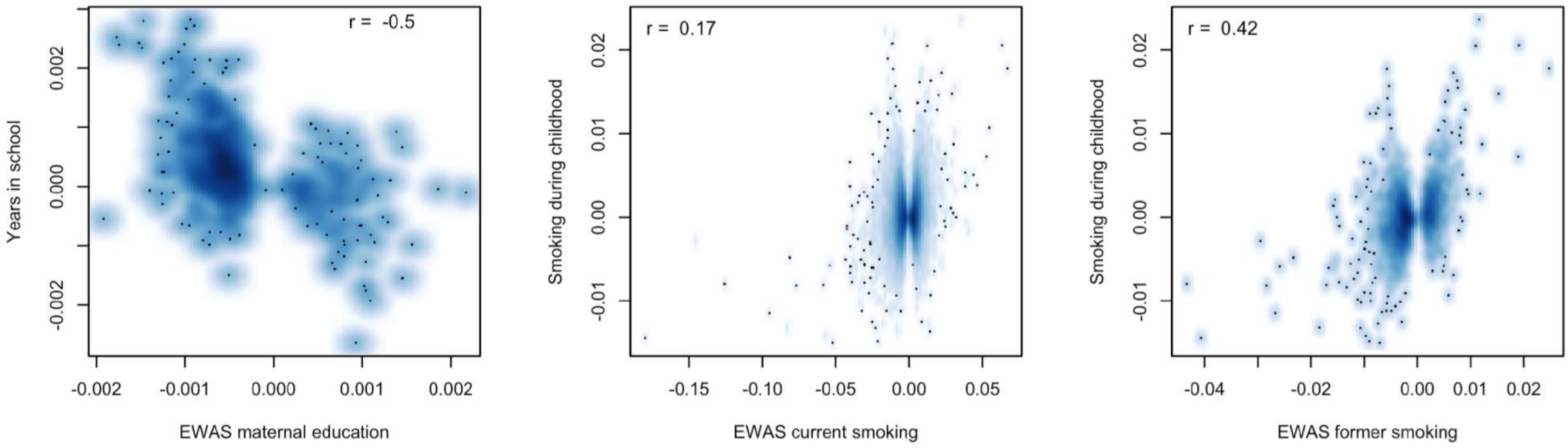


EWAS maternal education = Maternal educational attainment in pregnancy (PMID: 38052982)
EWAS current and former smoking = Epigenetic signatures of cigarette smoking (PMID: 27651444)

**Supplemental Table 10.** Replication results comparing single sites from the socioeconomic and behavior adjusted epigenome-wide association study showing the difference in percent methylation for each early life exposure on later life DNA methylation in the Health and Retirement Study to two prior meta-analyses

**File = Supplemental Table 10-Replication Compare Single Site Results.xlsx**

**Supplemental Table 11.** Gene Ontology pathway enrichment analysis results from our socioeconomic and behavior models for four early life exposures and later life DNA methylation in the Health and Retirement Study

**File = Supplemental Table 11-Gene Ontology Pathway Enrichment Results.xlsx**

**Supplemental Table 12.** Mixtures early life exposure regression results for age acceleration, global methylation, and site-specific methylation for socioeconomic and behavior adjusted models in the Health and Retirement Study

**File = Supplemental Table 12-Mixtures Early Life Exposure Results.xlsx**
